# A tumor focused approach to resolving the etiology of DNA mismatch repair deficient tumors classified as suspected Lynch syndrome

**DOI:** 10.1101/2023.02.27.23285541

**Authors:** Romy Walker, Khalid Mahmood, Jihoon E. Joo, Mark Clendenning, Peter Georgeson, Julia Como, Sharelle Joseland, Susan G. Preston, Yoland Antill, Rachel Austin, Alex Boussioutas, Michelle Bowman, Jo Burke, Ainsley Campbell, Simin Daneshvar, Emma Edwards, Margaret Gleeson, Annabel Goodwin, Marion T. Harris, Alex Henderson, Megan Higgins, John L. Hopper, Ryan A. Hutchinson, Emilia Ip, Joanne Isbister, Kais Kasem, Helen Marfan, Di Milnes, Annabelle Ng, Cassandra Nichols, Shona O’Connell, Nicholas Pachter, Bernard J. Pope, Nicola Poplawski, Abiramy Ragunathan, Courtney Smyth, Allan Spigelman, Kirsty Storey, Rachel Susman, Jessica A. Taylor, Linda Warwick, Mathilda Wilding, Rachel Williams, Aung K. Win, Michael D. Walsh, Finlay A. Macrae, Mark A. Jenkins, Christophe Rosty, Ingrid M. Winship, Daniel D. Buchanan, the Family Cancer Clinics of Australia

## Abstract

Routine screening of tumors for DNA mismatch repair (MMR) deficiency (dMMR) in colorectal (CRC), endometrial (EC) and sebaceous skin (SST) tumors leads to a significant proportion of unresolved cases classified as suspected Lynch syndrome (SLS). SLS cases (n=135) were recruited from Family Cancer Clinics across Australia and New Zealand. Targeted panel sequencing was performed on tumor (n=137; 80xCRCs, 33xECs and 24xSSTs) and matched blood-derived DNA to assess for microsatellite instability status, tumor mutation burden, COSMIC tumor mutational signatures and to identify germline and somatic MMR gene variants. MMR immunohistochemistry (IHC) and *MLH1* promoter methylation were repeated. In total, 86.9% of the 137 SLS tumors could be resolved into established subtypes. For 22.6% of these resolved SLS cases, primary *MLH1* epimutations (2.2%) as well as previously undetected germline MMR pathogenic variants (1.5%), tumor *MLH1* methylation (13.1%) or false positive dMMR IHC (5.8%) results were identified. Double somatic MMR gene mutations were the major cause of dMMR identified across each tumor type (73.9% of resolved cases, 64.2% overall, 70% of CRC, 45.5% of ECs and 70.8% of SSTs). The unresolved SLS tumors (13.1%) comprised tumors with only a single somatic (7.3%) or no somatic (5.8%) MMR gene mutations. A tumor-focused testing approach reclassified 86.9% of SLS into Lynch syndrome, sporadic dMMR or MMR-proficient cases. These findings support the incorporation of tumor sequencing and alternate *MLH1* methylation assays into clinical diagnostics to reduce the number of SLS patients and provide more appropriate surveillance and screening recommendations.

## Introduction

The current diagnostic strategy for identifying Lynch syndrome, the most common inherited cancer syndrome, as recommended by the National Comprehensive Cancer Network (www.nccn.org, last accessed date: November 8^th^, 2022) and the Evaluation of Genomic Applications in Practice and Prevention group (1), involves screening tumours for evidence of DNA mismatch repair (MMR)-deficiency (dMMR) via immunohistochemical staining for loss of expression of one or more of the MMR proteins (MMR IHC) and/or for microsatellite instability (MSI). Loss of MLH1/PMS2 protein expression necessitates testing for *MLH1* promoter methylation (or *BRAF* V600E in colorectal cancers (CRCs)) and if negative, germline MMR gene testing. For other patterns of loss of expression, germline MMR testing is undertaken. This approach, while effective at identifying people with Lynch syndrome, still results in a significant proportion of dMMR tumors without identified *MLH1* methylation or germline MMR pathogenic variant, referred to as Lynch-like or suspected Lynch syndrome (SLS) (2). A diagnosis of SLS presents challenges for the clinician with regards to recommendations for ongoing cancer risk management and for screening for first-degree relatives. For the patient, an SLS diagnosis results in variable psychosocial and behavioral responses related to the interpretation of their diagnosis (3, 4).

Previous studies have shown the SLS group to be etiologically heterogeneous, encompassing both inherited and sporadic causes of dMMR (2,5,6). Furthermore, the risk of cancer in SLS patients and their relatives requires clarification (2,7,8). These uncertainties make the clinical management of an SLS diagnosis challenging. Complex or cryptic germline MMR gene pathogenic variants that are more difficult to detect with current methodology, including those within intronic or regulatory regions, have been described (9–19). In addition, somatic mosaicism of MMR gene pathogenic variants (20, 21) or germline pathogenic variants in non-MMR genes, including *POLE*, *POLD1* or *MUTYH* that somatically inactivate one of the MMR genes (15, 22), are rare causes of tumor dMMR. The most commonly reported cause of SLS in CRC and endometrial cancer (EC) is biallelic somatic MMR gene mutations (often referred to as double MMR somatics) (23–27), where each of the two mutations inactivate an allele in the same MMR gene that is shown to be defective by the pattern of MMR protein loss of expression observed in the tumor. Biallelic somatic MMR gene mutations have also been reported in dMMR sebaceous skin tumors (SSTs) in the absence of germline MMR gene pathogenic variants (28). Furthermore, the possibility that an SLS diagnosis has arisen due to a false positive tumor MMR IHC result or false negative *MLH1* methylation test result has been previously described (25). The ability to stratify people with an SLS diagnosis into those with an incorrect screening test result or an inherited or sporadic etiology, is of clinical importance for risk appropriate clinical management of the patient and their relatives.

CRCs, ECs and SSTs are tumor types that demonstrate the highest frequencies of dMMR, where up to 26% (29), 31% (29, 30) and 31% (31) of these tumor types respectively, present with dMMR. The aim of this study was to investigate both inherited and somatic causes of 135 CRC-, EC-, or SST-affected people with an SLS diagnosis referred from Family Cancer Clinics across Australia and New Zealand. The findings from this large cohort with SLS will inform future diagnostic approaches that will improve the stratification of patients into those with a definite diagnosis of Lynch syndrome and those with somatic causes of dMMR. It will also eliminate the genetic counselling uncertainty of the finding of dMMR tumor where a somatic causation is demonstrable.

## Methods

### Study Cohort

The study participants were people diagnosed with SLS during clinical work-up. SLS was defined as: 1) having tumor dMMR as determined by MMR IHC where germline testing of the MMR genes did not find a pathogenic variant, 2) for tumors that showed loss of MLH1/PMS2 expression, tumor *MLH1* methylation testing returned a negative or inconclusive result, or 3) for CRC, where *MLH1* methylation testing was not completed, the tumor tested negative for the *BRAF* V600E mutation. Participants meeting the SLS criteria and with tumor tissue and blood-derived DNA available for testing were identified for this analysis. In total, 140 participants with SLS were identified for testing from two studies:

1) the ANGELS study (*<u>A</u>pplying <u>N</u>ovel <u>G</u>enomic approaches to <u>E</u>arly-onset and suspected <u>L</u>ynch <u>S</u>yndrome colorectal and endometrial cancers*) recruited SLS patients diagnosed with CRC and/or EC between 2014 – 2021 from Family Cancer Clinics across Australia and New Zealand (32) (n=124);
2) the <u>M</u>uir-<u>T</u>orre <u>S</u>yndrome Study (MTS) recruited people diagnosed with one or more SSTs between July 2016 and September 2021 from Sullivan Nicolaides Pathology in Brisbane, Australia (31) or from Family Cancer Clinics across Australia (n=26).

The study was approved by The University of Melbourne human research ethics committee (HREC#1750748 and HREC#1648355) and at certain Familial Cancer Clinic institutional review boards. All ANGELS and MTS study participants provided informed consent and a peripheral blood sample. Biopsy or resection tumor tissue blocks/slides were collected where possible.

### DNA Mismatch Repair Protein Immunohistochemistry

Pre-study MMR IHC testing to categorize the tumor as dMMR as part of the SLS diagnosis was performed by various diagnostic pathology services across Australia and New Zealand. For this study, MMR IHC was repeated as described in **Supplementary material** if tissue was available.

### Tumor MLH1 Methylation Testing

Pre-study tumor *MLH1* methylation testing was performed using the methylation sensitive- multiplex ligation probe dependent amplification (MS-MLPA) assay at various diagnostic pathology services across Australia. For this study, *MLH1* methylation testing employed a MethyLight assay (33, 34) and a methylation-sensitive high resolution melting assay (MS-HRM) (35), performed on the same tumor DNA sample from SLS cases that showed loss of MLH1/PMS2 expression or solitary loss of PMS2 expression (46, 47). These independent assays targeted seven overlapping CpG sites within the C-region of the *MLH1* gene promoter and were run with a set of DNA standards (0% - 100% methylation) and no-template (negative) controls. Bisulfite conversion of tumor and blood-derived DNA was performed using the EZ DNA Methylation- Lightning^TM^ Kit (Zymo Research, Irvine, USA). For MethyLight, *MLH1* methylation was quantitatively reported based on the percentage of methylated reference (PMR) calculations (34), where tumors with a PMR ≥10% were considered “positive” (33, 34). For MS-HRM, the MeltDoctor^TM^ HRM Reagent Kit (Thermo Fisher Scientific, Massachusetts, USA) was used where tumors demonstrating ≥5% were considered *MLH1* methylation “positive”. For each tumor positive for *MLH1* methylation, the matched blood-derived DNA sample was tested in people with tumors diagnosed <50 years or with multiple tumors using these two assays for evidence of constitutional *MLH1* methylation (*MLH1* epimutation).

### Targeted Multi-Gene Panel Testing

All tumors and matched blood-derived DNA samples from the n=135 SLS cases underwent multi- gene panel sequencing assay, modified from the assay described in Zaidi *et al.* (36), which captured 297 genes (2.005 megabases (Mb)). The panel comprised the MMR and *EPCAM* genes as well as other established hereditary CRC and EC genes including *POLE, POLD1,* and *MUTYH*. Details of the capture design and sequencing are provided in the **Supplementary Material.** Details of the bioinformatic pipeline for variant calling as well as methodology for calculation of tumor mutational burden (TMB) and tumor mutational signatures (TMS) are provided in the **Supplementary Material**.

### Determining Tumor DNA Mismatch Repair Deficiency from Panel Sequencing Data

Overall tumor dMMR status was determined from the panel sequencing data by applying the additive feature combination approach described in Walker *et al.*, (37) (**Supplementary Table S1)**. Briefly, six dMMR predictive features, namely MSMuTect, MANTIS, MSIseq, MSISensor, INDEL count and TMS ID2+ID7 (32) were derived for each tumor with thresholds for classifying dMMR determined previously (37) (see **Supplementary Table S1**). Tumors were considered dMMR overall when ≥3/6 of the features were positive for dMMR.

### Statistical Analysis

All statistical analyses were done using the R programming language (v. 4.1.0) (38). Correlation scores for categorical values between multiple groups were estimated using the *chi-square* test. *p*- values <0.05 were considered statistically significant.

## Results

### Characteristics of the SLS Study Participants

An overview of the study design is shown in **Figure 1** and includes the categorization of the SLS cases using the results from tumor sequencing as well as re-testing of *MLH1* methylation and MMR IHC which is described in detail below. The clinicopathological characteristics of the 137 tumors with sufficient DNA for testing from 135 study participants meeting the SLS criteria, overall and by tumor type, are presented in **Table 1**. Of note, two participants each had a CRC that showed loss of all four MMR proteins, where one tumor (ID018) was from a carrier of a germline *MSH2* pathogenic variant explaining the loss of MSH2/MSH6 expression but where the loss of MLH1/PMS2 expression was unexplained, while the other tumor (ID046) had no cause for loss of expression of all four MMR proteins during pre-study clinical investigations.

**Figure 1.**
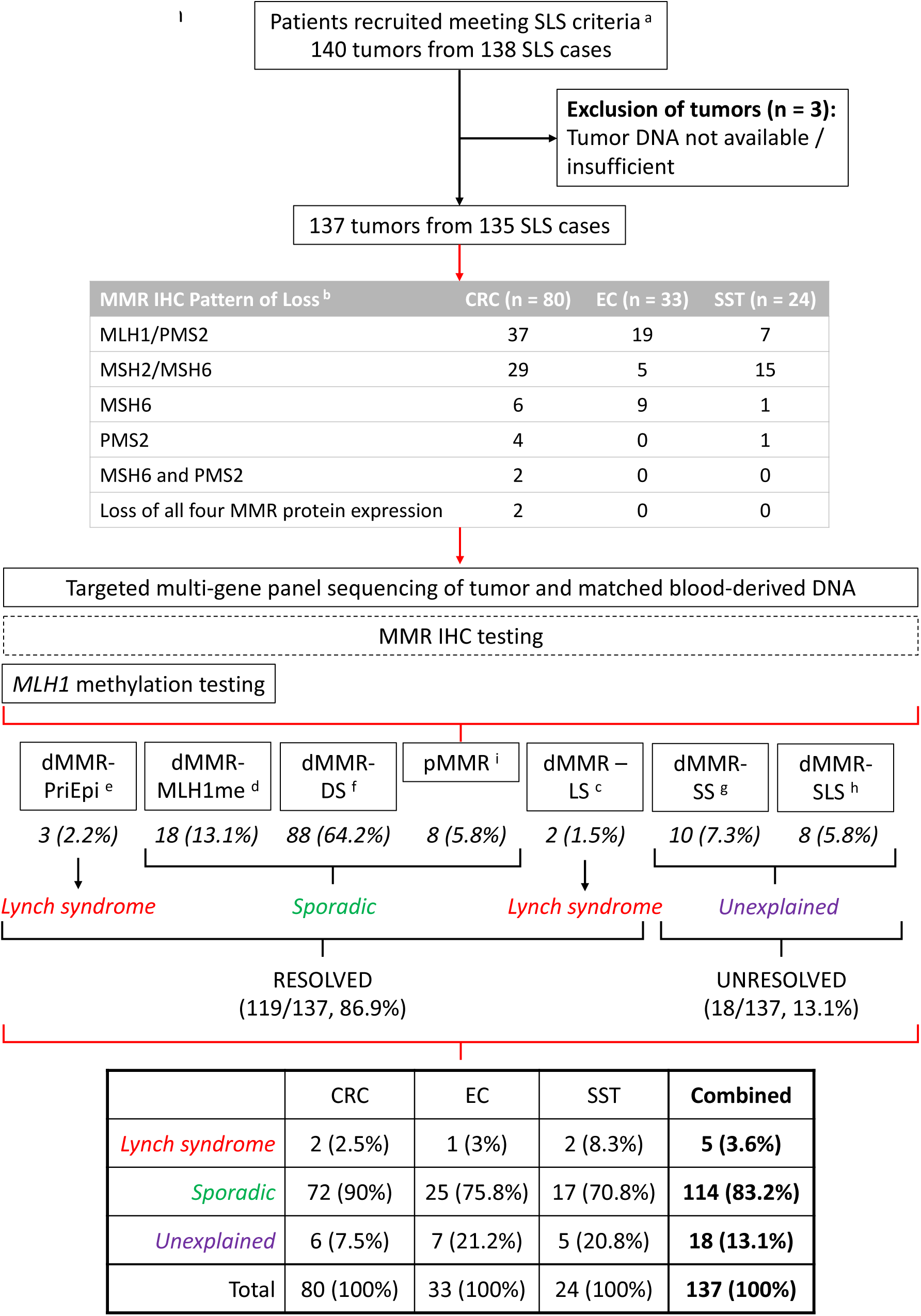
Overview of study design. Schema presenting the study inclusion criteria, the breakdown of the clinical MMR IHC results, the testing assays applied and the final study results, separated by tissue type and combined. Abbreviations: suspected Lynch syndrome, SLS; colorectal cancer, CRC; endometrial cancer, EC; sebaceous skin tumor, SST; DNA mismatch repair, MMR; immunohistochemistry, IHC; primary epimutation, dMMR-PriEpi; positive *MLH1* methylation, dMMR-MLH1me; double somatic mutations, dMMR-DS; DNA mismatch repair proficient, pMMR; Lynch syndrome, dMMR-LS; single somatic mutation, dMMR-SS. ^a^ SLS criteria: individuals diagnosed with a DNA mismatch repair deficient CRC, EC and/or SST with previous negative testing results. ^b^ Breakdown of clinical MMR IHC results when first entering the study. ^c^ dMMR with a germline pathogenic variant identified (Lynch syndrome, “dMMR-LS”) ^d^ dMMR with tumor MLH1 methylation (MLH1 methylated, “dMMR-MLH1me”) ^e^ dMMR with tumor and blood MLH1 methylation (primary epimutation, “dMMR-PriEpi”) ^f^ dMMR with double somatic MMR variants in the same MMR gene (double somatic mutation, “dMMR-DS”) ^g^ dMMR with a single somatic MMR variant (single somatic mutation, “dMMR-SS”) ^h^ dMMR with no germline or somatic variants (suspected Lynch syndrome, “dMMR-SLS”) ^i^ pMMR tumors with neither germline or somatic mutations nor hypermethylation of the MLH1 gene (DNA mismatch repair proficient, “pMMR”)

**Table 1.**
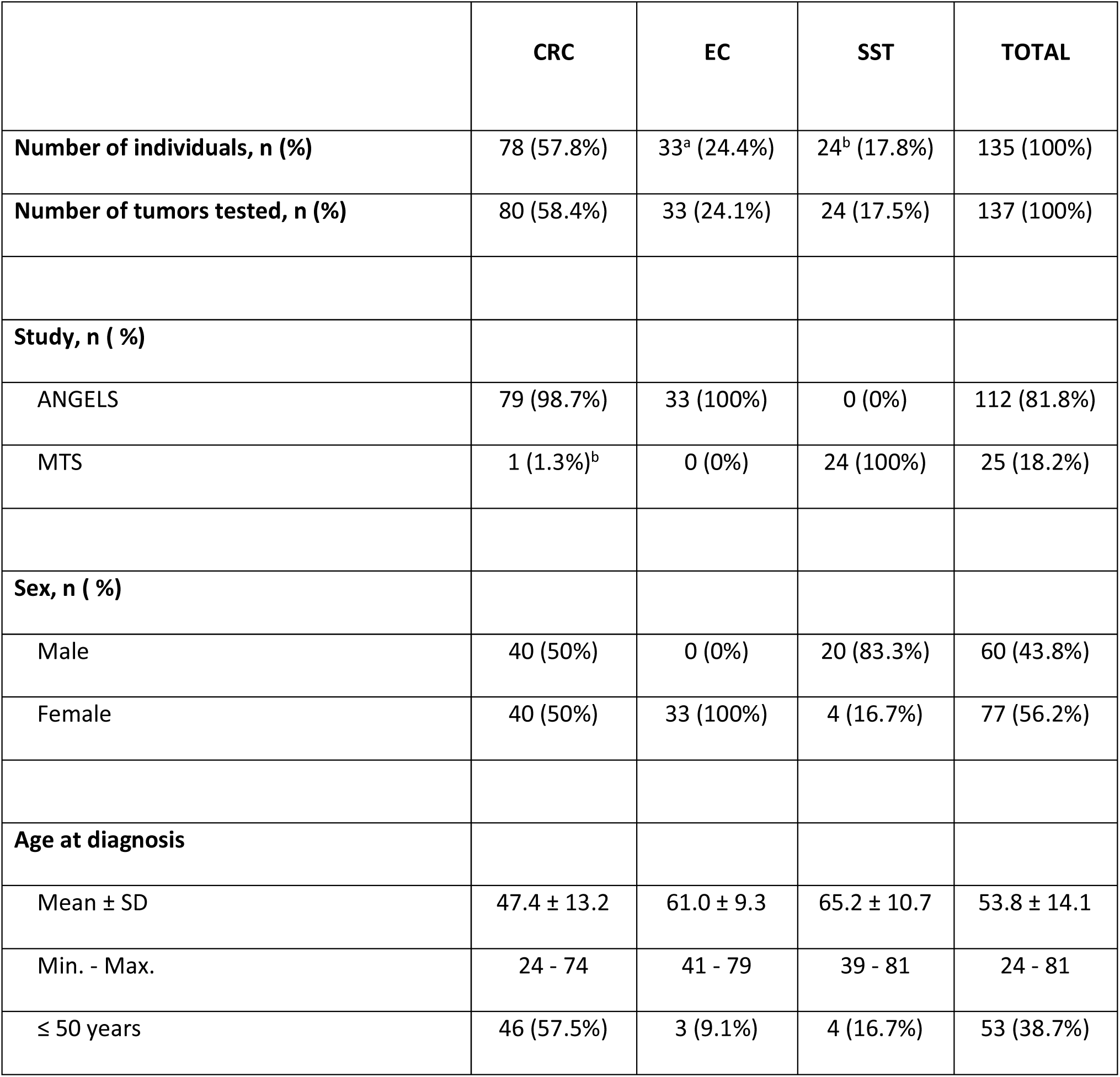
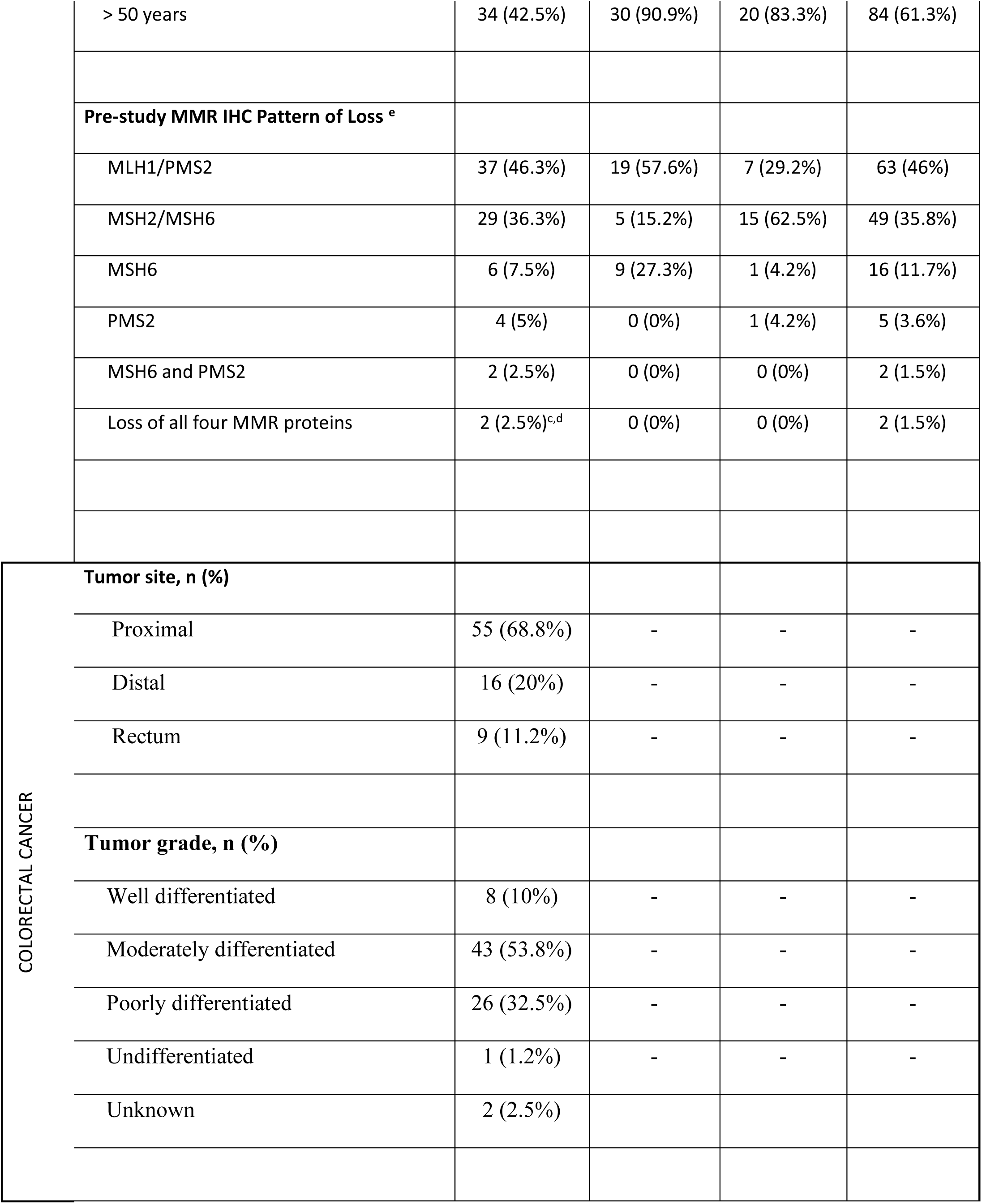
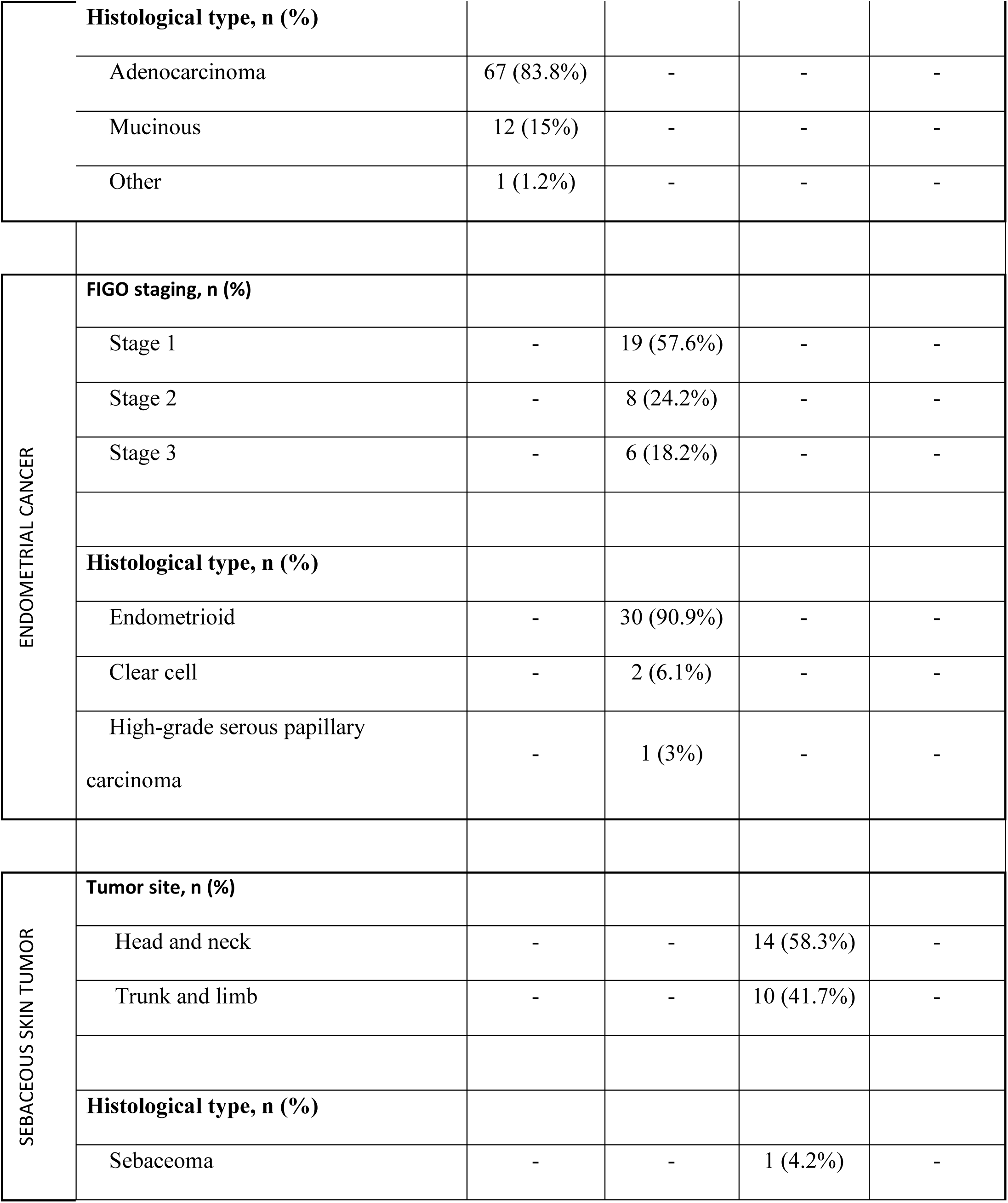
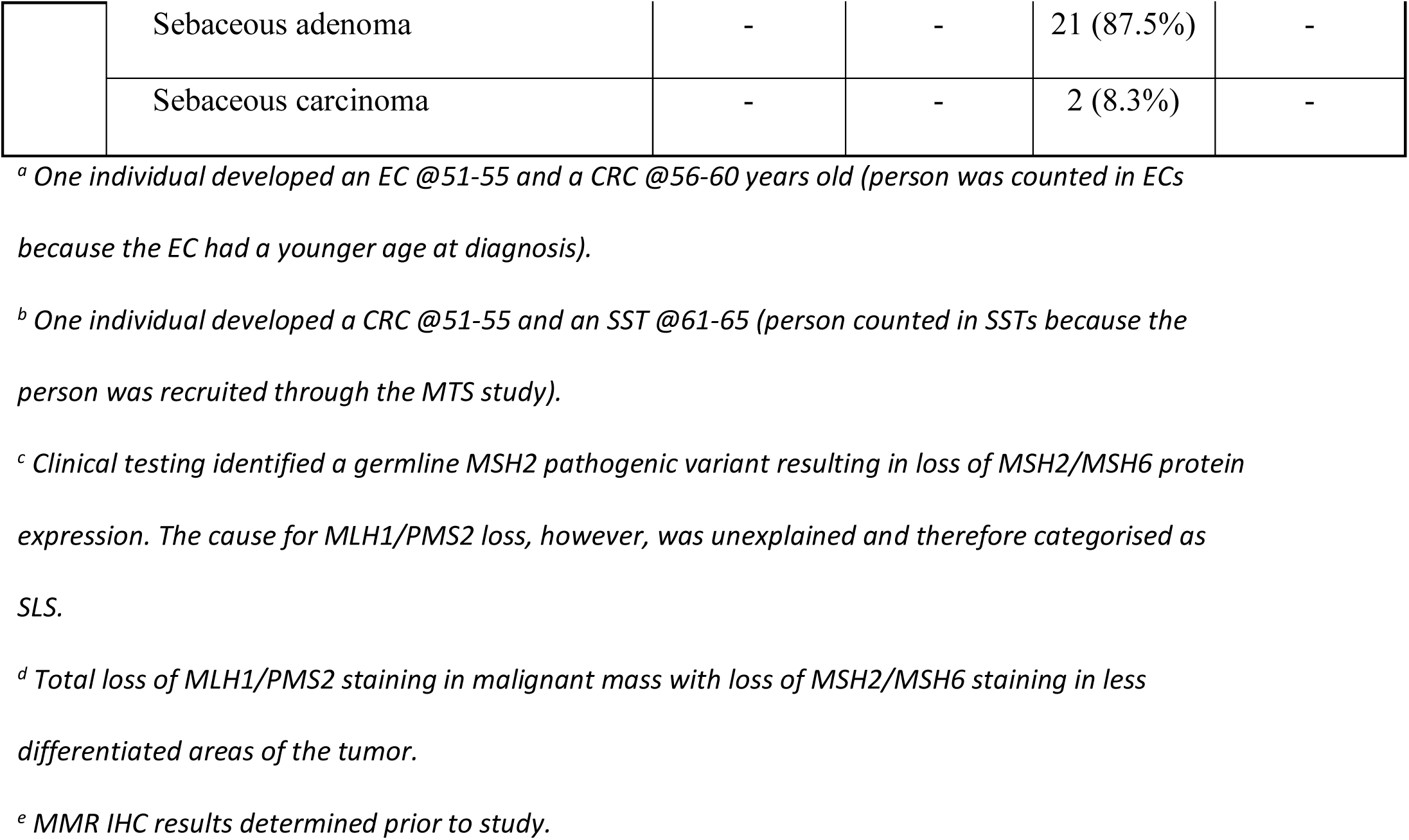
Overview of the study participants and their clinicopathological features overall and by tumor type. Abbreviations: colorectal cancer, CRC; endometrial cancer, EC; sebaceous skin tumor, SST; Applying Novel Genomic approaches to Early-onset and suspected Lynch Syndrome colorectal and endometrial cancers, ANGELS; Muir-Torre Syndrome, MTS; standard deviation, SD; Amsterdam II criteria, AM II; International Federation of Gynecology and Obstetrics, FIGO.

|  | CRC | EC | SST | TOTAL |
| --- | --- | --- | --- | --- |
| **Number of individuals, n (%)** | 78 (57.8%) | 33<sup>a</sup> (24.4%) | 24<sup>b</sup> (17.8%) | 135 (100%) |
| **Number of tumors tested, n (%)** | 80 (58.4%) | 33 (24.1%) | 24 (17.5%) | 137 (100%) |
| **Study, n ( %)** |  |  |  |  |
| ANGELS | 79 (98.7%) | 33 (100%) | 0 (0%) | 112 (81.8%) |
| MTS | 1 (1.3%)<sup>b</sup> | 0 (0%) | 24 (100%) | 25 (18.2%) |
| **Sex, n ( %)** |  |  |  |  |
| Male | 40 (50%) | 0 (0%) | 20 (83.3%) | 60 (43.8%) |
| Female | 40 (50%) | 33 (100%) | 4 (16.7%) | 77 (56.2%) |
| **Age at diagnosis** |  |  |  |  |
| Mean ± SD | 47.4 ± 13.2 | 61.0 ± 9.3 | 65.2 ± 10.7 | 53.8 ± 14.1 |
| Min. - Max. | 24 - 74 | 41 - 79 | 39 - 81 | 24 - 81 |
| ≤ 50 years | 46 (57.5%) | 3 (9.1%) | 4 (16.7%) | 53 (38.7%) |

|  |  |  |  |  |  |
| --- | --- | --- | --- | --- | --- |
|  | > 50 years | 34 (42.5%) | 30 (90.9%) | 20 (83.3%) | 84 (61.3%) |
|  | **Pre-study MMR IHC Pattern of Loss <sup>e</sup>** |  |  |  |  |
|  | MLH1/PMS2 | 37 (46.3%) | 19 (57.6%) | 7 (29.2%) | 63 (46%) |
|  | MSH2/MSH6 | 29 (36.3%) | 5 (15.2%) | 15 (62.5%) | 49 (35.8%) |
|  | MSH6 | 6 (7.5%) | 9 (27.3%) | 1 (4.2%) | 16 (11.7%) |
|  | PMS2 | 4 (5%) | 0 (0%) | 1 (4.2%) | 5 (3.6%) |
|  | MSH6 and PMS2 | 2 (2.5%) | 0 (0%) | 0 (0%) | 2 (1.5%) |
|  | Loss of all four MMR proteins | 2 (2.5%)<sup>c,d</sup> | 0 (0%) | 0 (0%) | 2 (1.5%) |
| COLORECTAL CANCER | **Tumor site, n (%)** |  |  |  |  |
|  | Proximal | 55 (68.8%) | - | - | - |
|  | Distal | 16 (20%) | - | - | - |
|  | Rectum | 9 (11.2%) | - | - | - |
|  | **Tumor grade, n (%)** |  |  |  |  |
|  | Well differentiated | 8 (10%) | - | - | - |
|  | Moderately differentiated | 43 (53.8%) | - | - | - |
|  | Poorly differentiated | 26 (32.5%) | - | - | - |
|  | Undifferentiated | 1 (1.2%) | - | - | - |
|  | Unknown | 2 (2.5%) |  |  |  |
|  | **Histological type, n (%)** |  |  |  |  |
|  | Adenocarcinoma | 67 (83.8%) | - | - | - |
|  | Mucinous | 12 (15%) | - | - | - |
|  | Other | 1 (1.2%) | - | - | - |
| ENDOMETRIAL CANCER | **FIGO staging, n (%)** |  |  |  |  |
|  | Stage 1 | - | 19 (57.6%) | - | - |
|  | Stage 2 | - | 8 (24.2%) | - | - |
|  | Stage 3 | - | 6 (18.2%) | - | - |
|  | **Histological type, n (%)** |  |  |  |  |
|  | Endometrioid | - | 30 (90.9%) | - | - |
|  | Clear cell | - | 2 (6.1%) | - | - |
|  | High-grade serous papillary carcinoma | - | 1 (3%) | - | - |
| SEBACEOUS SKIN TUMOR | **Tumor site, n (%)** |  |  |  |  |
|  | Head and neck | - | - | 14 (58.3%) | - |
|  | Trunk and limb | - | - | 10 (41.7%) | - |
|  | **Histological type, n (%)** |  |  |  |  |
|  | Sebaceoma | - | - | 1 (4.2%) | - |
|  | Sebaceous adenoma | - | - | 21 (87.5%) | - |
|  | Sebaceous carcinoma | - | - | 2 (8.3%) | - |

<sup>a</sup> One individual developed an EC @51-55 and a CRC @56-60 years old (person was counted in ECs
because the EC had a younger age at diagnosis).
<sup>b</sup> One individual developed a CRC @51-55 and an SST @61-65 (person counted in SSTs because the person was recruited through the MTS study).
<sup>c</sup> Clinical testing identified a germline MSH2 pathogenic variant resulting in loss of MSH2/MSH6 protein expression. The cause for MLH1/PMS2 loss, however, was unexplained and therefore categorised as SLS.
<sup>d</sup> Total loss of MLH1/PMS2 staining in malignant mass with loss of MSH2/MSH6 staining in less differentiated areas of the tumor.
<sup>e</sup> MMR IHC results determined prior to study.

### Determining Tumor dMMR Status

For the SLS tumors, firstly, confirmation of dMMR status was assessed using both the additive feature approach combining the results from MSMuTect, MANTIS, MSIseq, MSISensor, INDEL count and TMS ID2+ID7 as described in Walker *et al.*, (37) and by repeating MMR IHC where possible. The results of the additive feature approach, overall and for each tumor type, are shown in **Supplementary Figure S1**, where 85.4% (117/137) were predicted to be dMMR having ≥3/6 tumor features, including 87.5% (70/80) of the CRCs, 69.7% (23/33) of the ECs and all the SSTs (100%, 24/24). Of these 117 dMMR predicted tumors, 81.2% had all six tumor features positive for dMMR.

MMR IHC was repeated internally for 65/137 (47.4%) SLS tumors. Discordant MMR IHC results between the pattern of loss reported prior to the study entry compared with testing completed during the study were observed in 20% (13/65) of the SLS tumors (**Supplementary Table S2**). For 8/13 (61.5%) of these SLS tumors (7 CRCs and 1 EC) retained/normal expression of the MMR proteins was observed when repeated. All eight were predicted to be pMMR results by the additive feature combination approach. Furthermore, no tumor *MLH1* methylation or double somatic MMR mutations were identified in this group from internal testing, supporting a final categorization of pMMR. Five SLS tumors showed a different pattern of MMR protein loss compared with the pre-study result (5/13, 38.5%) (**Supplementary Table S2**). In each case, the new pattern of loss was consistent with cause of dMMR identified by this study. For example, ID009 showed solitary loss of MSH6 expression initially and when repeated internally showed loss of MLH1/PMS2 that was related to tumor *MLH1* methylation. There were 12 tumors that were classified as dMMR by MMR IHC but determined to be pMMR by the additive feature combination approach giving an overall accuracy between tumor panel sequencing derived dMMR status and the MMR IHC status of 92% (95% confidence intervals, CI: 86.5%-92%) (**Supplementary Table S3**).

### Evidence of Tumor MLH1 Methylation

The dual MethyLight and MS-HRM *MLH1* methylation assay approach was performed on 77 SLS tumors, including all 47 tumors which had pre-study clinical *MLH1* methylation testing. Tumor *MLH1* methylation was detected in 23 tumors from 22 SLS cases where the concordance between the two internal assays was 100% and, in all but one of the tumors, there was loss of expression of MLH1 protein by IHC (a single *MLH1* methylation positive tumor ID031 showed solitary loss of PMS2). Five of these tumors had pre-study clinical *MLH1* methylation testing reporting no *MLH1* methylation detected (4/5 were EC tumors) (**Supplementary Table S4**). There were six SLS tumors that reported inconclusive *MLH1* methylation results from pre-study clinical testing that were found to be positive for *MLH1* methylation, although at low levels, by this study (**Supplementary Table S4**). Two of the SLS cases were identified by the study as a primary *MLH1* epimutation carrier (ID033 and ID013; dMMR-PriEpi) showing *MLH1* methylation in their SST- and peripheral blood-derived DNA, and in the case of ID013 in their CRC tissue-derived DNA as well. Two SLS cases showed tumor *MLH1* methylation while also being a carrier of a germline MMR pathogenic variant (ID018 and ID034) demonstrating two concurrent mechanisms that accounted for the unique patterns of MMR protein loss observed in both (**Supplementary Table S4**). Therefore, 18/23 *MLH1* methylation positive tumors were re-categorized from SLS to sporadic *MLH1* methylated tumors (dMMR-MLH1me). Of all the *MLH1* methylation positive cases identified in this study, 55.6% (5/9) of the CRCs were diagnosed ≤50 years of age, whereas all *MLH1* methylation positive ECs (n=9) were diagnosis >50 years of age.

### Determining a Germline Cause of dMMR in SLS

The germline pathogenic variants and variants of uncertain significance (VUS) identified in the DNA MMR genes, *MUTYH*, and the exonuclease domain of *POLE* genes are shown in **Supplementary Table S4**. There were no germline pathogenic variants or VUS’s identified inside the exonuclease domain of the *POLD1* gene. Two germline MMR gene pathogenic variant carriers were identified (dMMR-LS). One, an *MSH2* deletion of exon 7 was known prior to study entry (ID018) with the CRC tumor showing loss of all four MMR proteins and was positive for *MLH1* methylation. The second carrier (*MSH6* c.3834_3849dup p.Thr1284Glnfs*10) was identified in ID034 who had MLH1/PMS2 and MSH6 loss in EC diagnosed at 55-60 years that was not reported in previous clinical testing. The tumor showed a somatic *MSH6* mutation (*MSH6*: c.3261del p.Phe1088Serfs*2) and was positive for *MLH1* methylation accounting for the loss of MLH1/PMS2. The third case harbored an *MLH1* VUS (*MLH1* c.400A>G p.Lys134Glu in ID028) identified in an SST tumor showing loss of MLH1/PMS2 and two somatic *MLH1* mutations. A further six VUS variants were identified in MMR genes which did not match the defective MMR gene identified by the pattern of MMR IHC loss. No biallelic *MUTYH* carriers were identified. However, two germline *POLE* variants within the exonuclease domain were observed, c.825C>G p.Asp275Glu and c.861T>A p.Asp287Glu, both of which are considered to be VUS (**Supplementary Table S5)**.

### Determining Double Somatic MMR Mutations as a Cause of dMMR in SLS

For the remaining 105 tumors not categorized as pMMR, dMMR-MLH1me, dMMR-PriEpi or dMMR-LS, two somatic mutations in the MMR gene indicated to be defective by the pattern of MMR IHC loss were identified in 87/105 (82.9%) tumors (**Table 2**). The presence of two or more somatic MMR mutations in each tumor was specific to the double somatic MMR mutations (dMMR-DS) tumors compared with the other tumor subtypes (**Supplementary Figure S2**). The somatic mutations comprised either two single nucleotide/small indel mutations or a single nucleotide/small indel mutation combined with a large deletion in the wildtype allele (loss of heterozygosity, LOH) (**Supplementary Figure S3**). When the tumors were stratified by their revised pattern of protein loss by IHC, >80% of tumors for each pattern were dMMR-DS across all tumor types (**Table 2**). Single somatic MMR gene mutations (dMMR-SS) occurred in 9.5% of the SLS tumors while no somatic MMR mutations (dMMR-SLS) were found in 7.6% (**Table 2**).

**Table 2.**
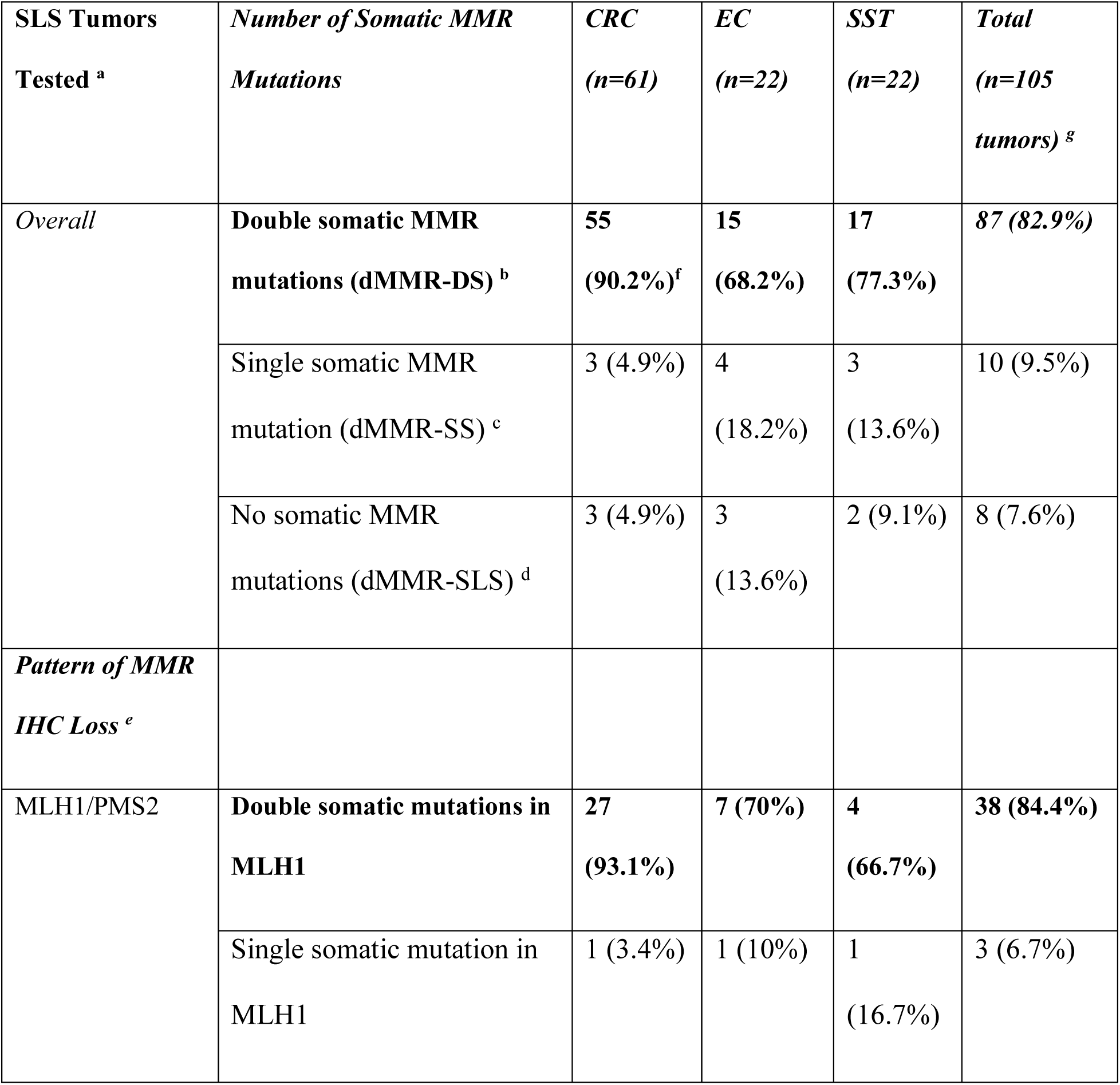
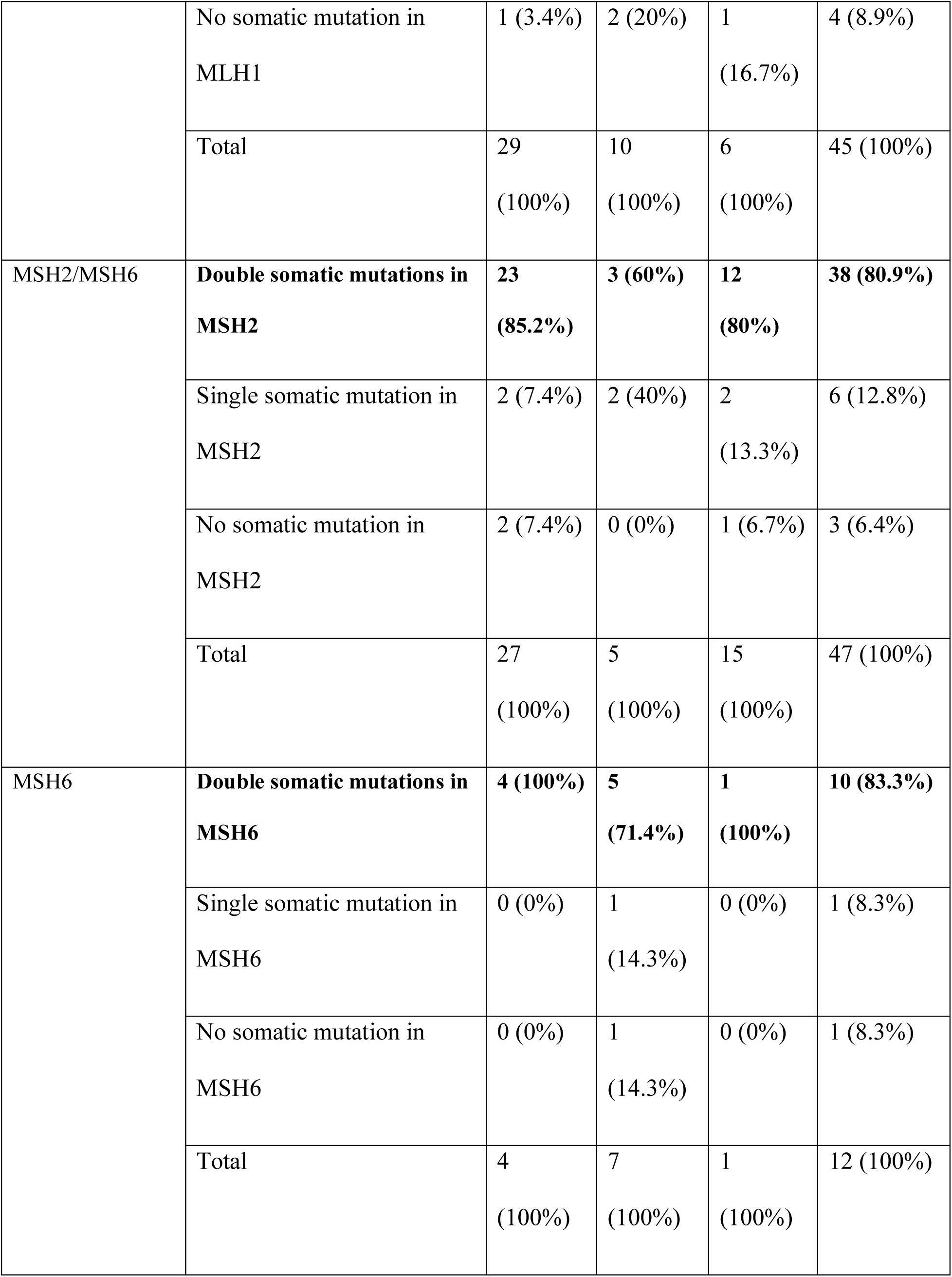
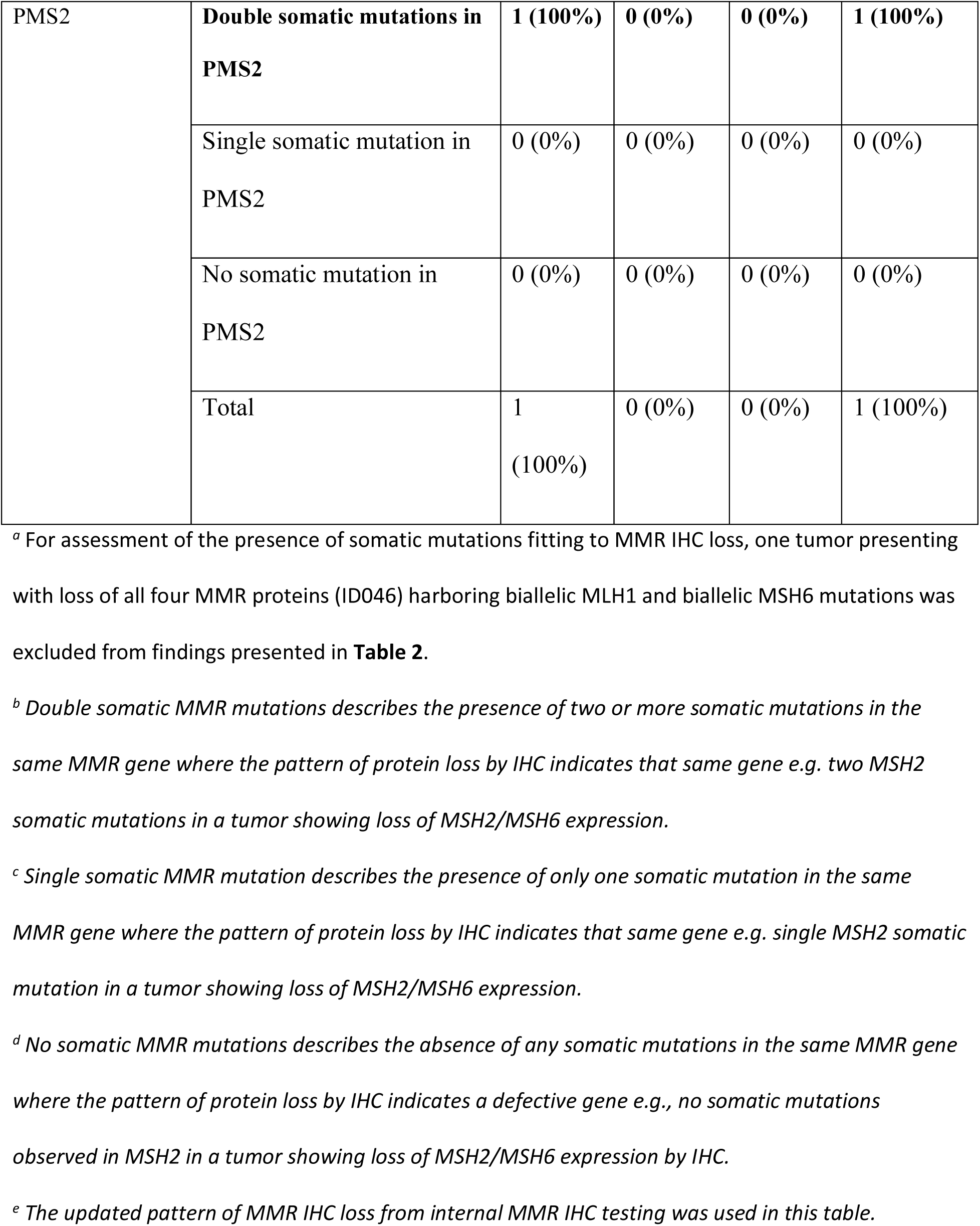
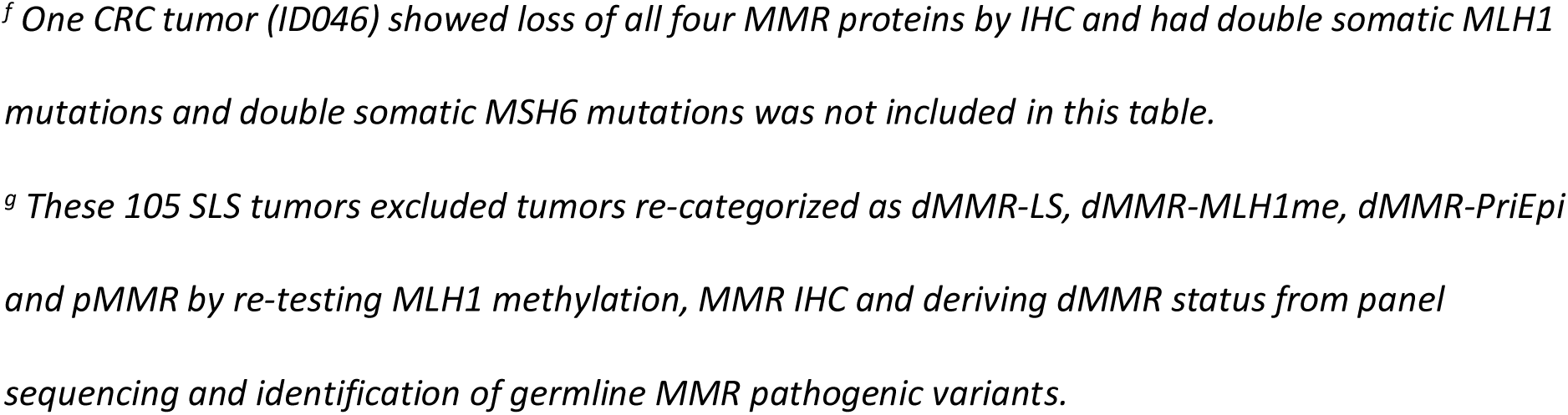
Overview of somatic mutation count by tumor type and by observed MMR IHC. Abbreviations: colorectal cancer, CRC; endometrial cancer, EC; sebaceous skin tumor, SST; immunohistochemistry, IHC; DNA mismatch repair, MMR; suspected Lynch syndrome, dMMR- SLS; DNA mismatch repair deficient tumor with double somatic mutations, dMMR-DS; DNA mismatch repair deficient tumor with single somatic mutation, dMMR-SS, DNA mismatch repair deficient tumor with no somatic mutations, dMMR-SLS.

For the dMMR-DS tumors, it was not possible to determine whether the double somatic mutations in the same MMR gene were in *cis* or *trans*. To address this, the number of somatic MMR mutations identified in each tumor across all four MMR genes were mapped to the pattern of MMR protein loss by IHC (**Figure 2**). Two or more somatic MMR mutations were rarely found in an MMR gene not considered to have the primary defect by IHC. For example, in tumors that showed loss of MLH1/PMS2 expression, multiple somatic mutations were observed in *MLH1* but rarely in the *MSH2*, *MSH6* or *PMS2* genes (**Figure 2A**), suggesting that when multiple mutations occur in the gene with loss of expression, they are acting in *trans* to inactivate both alleles. Multiple somatic MMR mutations rarely occurred in the dMMR-MLH1me or pMMR tumors (**Figure 2B & 2C**).

**Figure 2:**
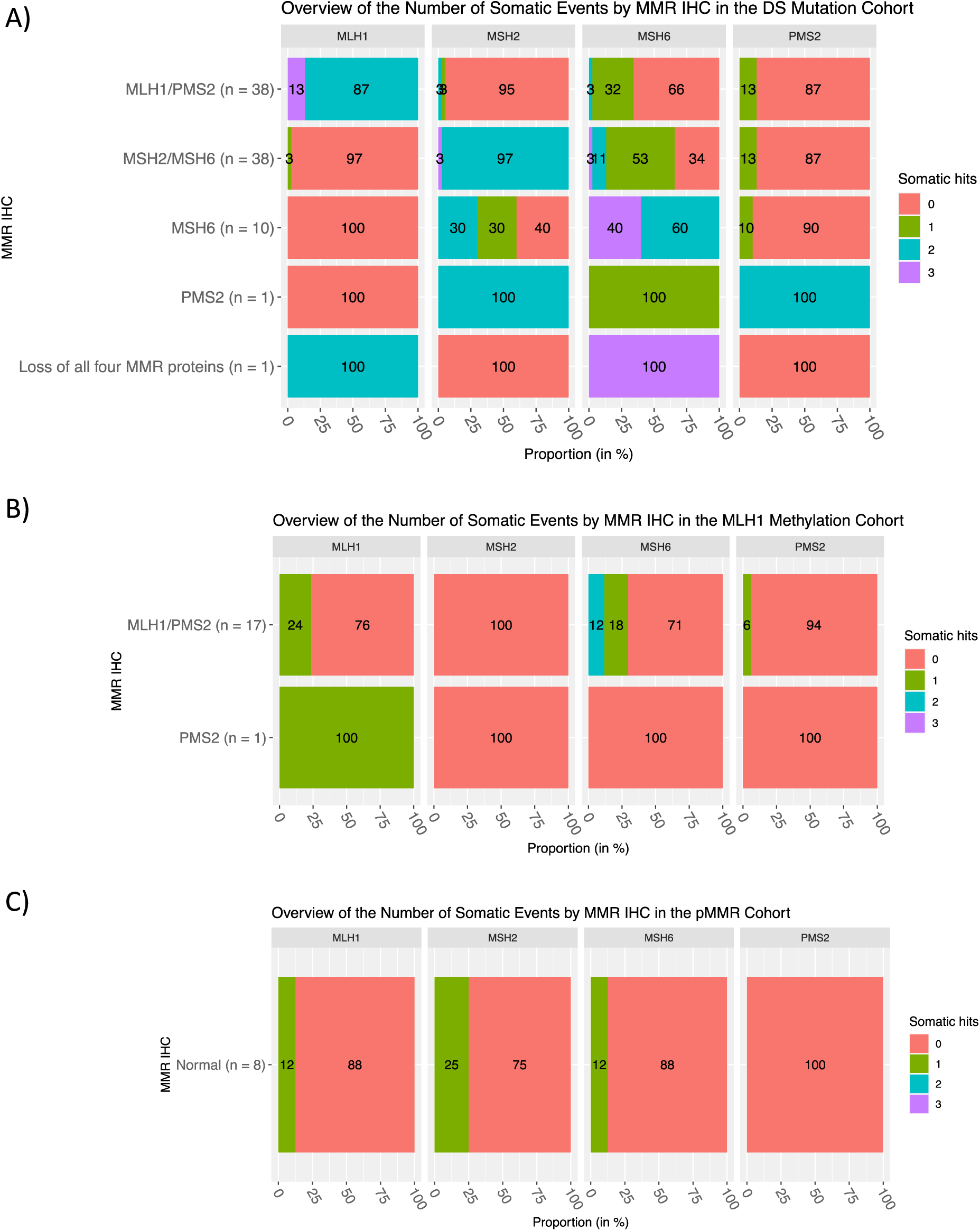
Overview of the number of somatic events (somatic mutation and LOH) by MMR IHC in the A) double somatic and B) positive *MLH1* methylation cohorts. Abbreviations: DNA mismatch repair, MMR; immunohistochemistry, IHC.

**Table 3** provides a summary of the categorization of all 137 SLS tumors overall and by tumor type. The cause for the dMMR phenotype, whether related to incorrect pre-study MMR IHC or *MLH1* methylation test result or identified germline or somatic cause, could be identified in 119/137 (86.9%) of the SLS cases and, therefore, considered resolved. The SLS tumors that were considered unresolved in terms of their dMMR etiology were those classified as dMMR-SS (7.3%, 10/137) and dMMR-SLS (5.8%, 8/137) (**Table 3**).

**Table 3.** Summary of the categorization of the SLS tumors, overall and by tumor type, based on the results from tumor panel sequencing, MLH1 methylation and DNA mismatch repair (MMR) immunohistochemistry (IHC) results. Table shows breakdown of sequencing results by tissue type and by cancer subtype. Abbreviations: colorectal cancer, CRC; endometrial cancer, EC; sebaceous skin tumor, SST; suspected Lynch syndrome, dMMR-SLS; DNA mismatch repair, MMR; DNA mismatch repair deficient, dMMR; DNA mismatch repair proficient, pMMR; DNA mismatch repair deficient tumor with double somatic mutations, dMMR-DS; DNA mismatch repair deficient tumor presenting with MLH1 methylation, dMMR-MLH1me; DNA mismatch repair deficient tumor with a primary MLH1 epimutation, dMMR-PriEpi; DNA mismatch repair deficient tumor with a germline pathogenic variant, dMMR-LS; DNA mismatch repair deficient tumor with single somatic mutation, dMMR-SS, DNA mismatch repair deficient tumor with no somatic mutations, dMMR-SLS.

| Category | CRC | EC | SST | Total |
| --- | --- | --- | --- | --- |
| Total Tumors Tested | 80 | 33 | 24 | 137 |
| ***Resolved*** |  |  |  |  |
| dMMR-DS - Double somatic MMR mutations | 56 (70%)<sup>a</sup> | 15 (45.5%) | 17 (70.8%) | 88 (64.2%) |
| dMMR-MLH1me - *MLH1* methylated<sup>b</sup> | 9 (11.3%) | 9 (27.3%) | 0 (0%) | 18 (13.1%) |
| dMMR-PriEpi - Primary *MLH1* epimutation | 1 (1.2%) | 0 (0%) | 2 (8.3%) | 3 (2.2%)<sup>e</sup> |
| dMMR-LS - Lynch syndrome | 1 (1.2%)<sup>c</sup> | 1 (3%)<sup>d</sup> | 0 (0%) | 2 (1.5%) |
| pMMR - MMR-proficient | 7 (8.8%) | 1 (3%) | 0 (0%) | 8 (5.8%) |
| **Total resolved** | **74/80**<br>**(92.5%)** | **26/33**<br>**(78.8%)** | **19/24**<br>**(79.2%)** | **119/137**<br>**(86.9%)** |
| ***Unresolved*** |  |  |  |  |
| dMMR-SS - Single somatic mutation | 3 (3.8%) | 4 (12.1%) | 3 (12.5%) | 10 (7.3%) |
| dMMR-SLS - remain as SLS | 3 (3.8%) | 3 (9.1%) | 2 (8.3%) | 8 (5.8%) |

<sup>a</sup> One tumor carried double somatic mutations in MLH1 and double somatic mutations in MSH6 presenting with loss of all four MMR proteins by IHC.
<sup>b</sup> All tumors positive for tumor MLH1 methylation demonstrated loss of MLH1/PMS2 by MMR IHC except for one CRC tumor showing solitary PMS2 loss by MMR IHC (confirmed by internal testing).
<sup>c</sup> This person carried a germline pathogenic variant in MSH2 (known prior to entering the study) with a somatic MSH2 mutation (2nd hit) and was also positive for tumor MLH1 methylation which accounted for the loss of all four MMR protein expression by IHC.
<sup>d</sup> This person carried a germline pathogenic variant in MSH6 (missed by prior clinical testing) with a somatic MSH6 mutation (2nd hit) as well as presenting with tumor MLH1 methylation accounting for the observed pattern of loss MLH1/PMS2 and MSH6 by MMR IHC.
<sup>e</sup> Two people identified as MLH1 epimutation carriers with one carrier developing an SST and the other developed a CRC and an SST.

### Characteristics of the dMMR-DS Tumors

The characteristics of the participants with dMMR-DS tumors including the sex, age at tumor diagnosis, PREMM5 scores and tumor site are shown in **Supplementary Table S6**. Two-thirds of the CRC dMMR-DS tumors were in the proximal colon (**Supplementary Table S6, Supplementary Figure S4**). The mean age at CRC diagnosis was 46.6 ± 13.1 years with 50% of the tumors diagnosed before age 50 years, in contrast to the EC and SST dMMR-DS tumors had an older mean age at diagnosis (**Supplementary Table S6**). The dMMR-DS CRCs located in the proximal colon had an older age at diagnosis compared with the dMMR-DS distal CRCs (p- value=0.043, *t-test*; **Supplementary Figure S5**). A PREMM5 score was calculated on each of the dMMR-DS categorized participants with the distribution of scores overall and by tumor type shown in **Supplementary Figure S6**. Over 80% of the dMMR-DS CRCs had a PREMM5 score greater than the 2.5 threshold, however, this proportion was much lower for the EC and SST groups (**Supplementary Table S6**).

## Discussion

In this study, we investigated both germline and somatic causes of dMMR using a custom- designed, multi-gene panel sequencing assay, and additionally investigated the potential of incorrect MMR IHC and tumor *MLH1* methylation results, in a large series of people diagnosed with SLS across CRC, EC and SST tumor types. Using this approach, we could resolve the diagnosis for 86.9% of the SLS tumors into recognized clinically actionable subtypes. The largest subtype of SLS tumors were those with double somatic MMR mutations (dMMR-DS, 64.2%) that are thought to be related to a low risk of second primary cancers and a low risk of cancer in relatives. Furthermore, 13.1% and 5.8% of SLS tumors were related to incorrect *MLH1* methylation and MMR IHC results, respectively, during pre-study clinical work-up. These results provide an important evidence base to improve tumor testing approaches for Lynch syndrome. Furthermore, our results highlight the added benefit to resolving an SLS diagnosis from deriving dMMR-associated features and tumor mutational signatures from tumor sequencing assay to confirm dMMR status and provide insights into tumor etiology.

The predominant cause of dMMR in the SLS CRC, EC and SST tumors was double somatic MMR mutations, resulting in somatic biallelic inactivation of the MMR gene, which is reflected in the pattern of protein loss identified by MMR IHC. After excluding tumors incorrectly categorized as SLS, 90.2% of CRCs, 68.2% of ECs and 77.3% of SSTs were identified as dMMR-DS (**Table 2**). Previous studies investigating SLS dMMR CRC and EC tumors have reported similarly high proportions with double somatic MMR mutations ranging from 52.5%-100% (5,6,23–26,39–41). Elze *et al.* (23), reported 88.8% (182/205) of dMMR CRCs and 80.9% (38/47) of dMMR ECs with two somatic inactivating events. Pearlman *et al.* (40) and Hampel *et al.* (24) identified double somatic MMR mutations in 88.4% (76/86) of dMMR SLS CRCs and in all of the 12 SLS ECs tested in the Ohio Colorectal Cancer Prevention Initiative study, respectively. For SSTs, Joly *et al.* (28) reported 53.8% (7/13) of the dMMR SLS tumors tested had likely double somatic MMR mutations. A study by Lefol *et al.* (6) investigated the prevalence of double somatic MMR mutations in multiple tumor types including CRC, EC and SST tumors observing 69.6%, 65% and 50%, respectively. Our study adds further confirmation that double somatic MMR mutations underlie the majority of the SLS dMMR subtype and supports the importance of incorporating tumor sequencing to resolve an SLS diagnosis. Furthermore, we have screened the largest group of SLS SSTs to date, demonstrating that double somatic MMR mutations are the most likely cause for dMMR after exclusion of Lynch syndrome.

The identification of only a single germline MMR pathogenic variant in *MSH6* that was missed by previous clinical germline testing was reassuring. Arnold *et al.* (9) reported 7% (9/128) of SLS cases had germline pathogenic variants identified that were missed by prior testing. The hotspot *MSH2* c.942+3A>T pathogenic variant (42) can be missed because it resides within a low DNA complexity region. The *MLH1* c.400A>G p.Lys134Glu VUS identified, occurred in a tumor with loss of MLH1/PMS2 expression and with two somatic *MLH1* mutations, where one of these may function as the “second hit” on the wildtype allele, however, further characterization of this variant is needed to determine whether this person has Lynch syndrome or double somatic MMR mutation-related dMMR. The other six MMR VUS occurred in genes that did not match the pattern of protein loss by MMR IHC and, therefore, this reduces their likelihood of being pathogenic. In addition, we investigated germline pathogenic variants in non-MMR genes namely, *MUTYH*, *POLE* and *POLD1*, as these have been previously shown to result in a double MMR somatic mutation dMMR phenotype (15, 22). We did not find germline biallelic *MUTYH* pathogenic variants nor did we see strong evidence for the tumor mutational signature profile, SBS18 and SBS36, that is strongly associated with germline biallelic inactivation of *MUTYH* gene (43) in any of the SLS tumors suggesting biallelic *MUTYH* inactivation is a rare cause of dMMR in SLS. Although we found only a single germline MMR and no non-MMR pathogenic variants in our SLS cases, the presence of a personal and/or family cancer history of Lynch syndrome spectrum tumors may provide cause for further investigation of these genes with alternate technology such as whole genome sequencing (41) or long-read genome sequencing (18), which have had success at identifying structural rearrangements and intronic pathogenic variants in the MMR genes.

Our approach to re-test tumor *MLH1* methylation and MMR IHC resulted in the identification of 18.9% of cases incorrectly classified as SLS, being either *MLH1* methylation positive tumors or being pMMR tumors. The study by Pearlman et al (27) found 13.7% of non-methylated CRCs had an incorrect MMR IHC result. There are recognized challenges with MMR IHC testing due to technical artefacts and inherent variability in the interpretation of the staining by different pathologists (44, 45). The pre-study MMR IHC was performed at multiple different private and public pathology laboratories across the country which may have led to the false positive IHC results we observed. The addition of our additive feature combination approach for predicting dMMR status from tumor sequencing data supported the reclassification of IHC results to pMMR in all eight cases. This highlights the value in applying alternate methodologies to confirm dMMR status when a diagnosis of SLS is made. Different patterns of loss were also observed in five SLS tumors, including four indicating loss of MLH1 which resulted in a further four SLS cases being tested for *MLH1* methylation, two of which were positive.

In addition to the false positive MMR IHC results, our study found 13.1% of the SLS tumors were indeed positive for tumor *MLH1* methylation indicating a large proportion was missed by pre-study clinical testing, particularly for the EC tumor type, which resulted in an incorrect SLS diagnosis. Of note, one SLS case with solitary loss of PMS2 expression was positive for *MLH1* methylation. *MHL1* methylation in tumors showing solitary PMS2 loss have been described previously (46, 47). Although the reason for these false negative results is difficult to definitively determine, potential reasons include: 1) intratumoral heterogeneity of *MLH1* methylation where different areas of the tumor were tested by the pathology labs and by the study, and 2) the sensitivity of *MLH1* methylation detection is likely different between different assays. The 100% concordance between the MethyLight and MS-HRM assay results while reassuring, also suggests these two assays may have increased sensitivity over MS-MLPA. This may be in part related to methodological differences relating to the need for bisulfite conversion for the MethyLight and MS-HRM assays compared with methylation-sensitive restriction enzyme for MS-MLPA. Our findings support the use of an alternate *MLH1* methylation assay when an SLS case with loss of MLH1/PMS2 is identified. A recent study that integrated *MLH1* methylation and targeted tumor sequencing is a promising approach to triage for Lynch syndrome where a single test would be more efficient and perhaps overcome some of the limitations of current MMR IHC and *MLH1* methylation testing (48).

Defective MMR gene function and loss of protein expression relies on the two-hit hypothesis requiring both alleles to be inactivated to drive tumorigenesis. The identification in our study, and reported in other studies using tumor sequencing to resolve SLS (6,25,28), that identification of only a single somatic MMR mutation presents a conundrum to the interpretation of dMMR etiology. The possibility that there is a second somatic mutation that has not been identified by our experimental approach e.g., intronic somatic mutation, or that there is an undetected germline MMR pathogenic variant (18, 25), is plausible given the dMMR tumor status, although each would have a different outcome for clinical management. The observation in this study that single somatic MMR mutations occurred in MMR genes not considered defective by the pattern of protein loss by IHC (**Figure 2**) and that single somatic MMR mutations occurred in *MLH1* methylation positive tumors and even in pMMR tumors (**Supplementary Figure S2**) suggests a single somatic MMR mutation can occur unrelated to the dMMR etiology, hence our categorization of the dMMR-SS tumors as unresolved.

The strengths of this study include the large number of cases diagnosed with SLS based on prior clinical work-up identified from family cancer clinics across each state of Australia and from New Zealand, representing the real-world heterogeneity of cases, diagnostic laboratory methodology and nuanced approaches to triaging for Lynch syndrome. Furthermore, tumor types representing those with the highest prevalence of dMMR, CRC, EC, and SST, were studied where the diagnosis of SLS is more likely to occur. The decision to repeat *MLH1* methylation and MMR IHC testing with different methodology resolved a larger number of SLS cases. Our custom-designed tumor sequencing assay enabled the investigation of multiple causes of dMMR simultaneously including SLS cases with unusual patterns of protein loss by IHC, including an SLS case with loss of all four MMR proteins that harbored double somatic mutations in *MLH1* and in *MSH6*. Furthermore, evaluation of multiple NGS-derived tumor features namely TMB, INDEL count, multiple MSI calling tools and COSMIC TMS enabled accurate dMMR prediction to support the MMR IHC result. Lastly, screening for *MLH1* epimutations in blood-derived DNA in SLS tumors with loss of MLH1/PMS2 diagnosed <50 years and in all six SST tumors with loss of MLH1/PMS2 identified two primary epimutation carriers, both in SST.

The identification of double somatic MMR mutations implies the dMMR tumor has a sporadic etiology, however, there remains some uncertainty that this is truly the case. This is in part due to previous reports showing that in rare cases a germline MMR pathogenic variant that is difficult to detect with current sequencing technology, including intronic pathogenic variants or a cryptic or complex germline variant may underlie the dMMR tumor phenotype (9,10,17,18,49). Although our capture was designed to include probes to cover non-coding regions of the MMR genes, not all these regions could be probed due to low sequencing complexity. We have previously tried to address the idea of missing intronic and complex MMR pathogenic variants using whole genome sequencing but found no viable germline MMR gene candidates in familial and/or early-onset SLS cases (41). Furthermore, this study did not include screening for potential somatic mosaicism of MMR variants in the dMMR-DS group, which would require deep sequencing analysis to detect low level mosaic mutations and screening of other distinct DNA sources. Somatic MMR mosaicism has been previously described (20, 21) although is rare. Follow-up studies of this potential mechanism are needed as the identification of post-zygotic mosaicism of an MMR pathogenic variant would have implications for future cancer risk and potentially for the carrier’s offspring. The unresolved group dMMR-SS and dMMR-SLS tumors, comprising 13.5% of the SLS tumors, remain categorized as SLS and will require further investigation to determine a somatic, germline or technical cause for their dMMR tumor. Finally, we were not able to investigate the original MMR IHC result/slides for the 20% of tumors that were identified as misclassified and, therefore, could not determine the basis, whether technical or from staining interpretation, for the pre-study MMR IHC result. Further engagement of quality assurance programs for MMR IHC and training for Pathologists may be needed to minimize the number of false positive / negative MMR IHC results and to trigger further laboratory investigations before reporting when unusual patterns of loss e.g. MSH6 and PMS2 loss are observed, as was reported pre-study for two SLS cases in this study.

## Conclusion

This study demonstrated a tumor-focused approach that incorporated multiple pieces of evidence, including contemporary NGS-derived tumor features and somatic screening of the MMR genes to resolve 86.9% of the SLS cases into clinically actionable subtypes. These findings provide an evidence base to reduce the number of patients diagnosed with SLS and improve triaging for Lynch syndrome. The increased implementation of tumor sequencing to identify double somatic MMR mutations will improve risk appropriate clinical management of the patient and their relatives. Further studies are needed to elucidate the non-coding regions of the MMR genes and to clarify the cancer risks for first degree relatives associated with people with double somatic MMR mutation tumor as currently the evidence is limited and focused on the heterogeneous SLS subtype (7). A large and systematic study of somatic mosaicism is needed in double somatic MMR mutation tumors to understand the true prevalence. Finally, efficient triage of cancer-affected people for Lynch syndrome should start with tumor and matched germline sequencing of the MMR genes (among others), for the determination of dMMR status, identification of double somatic MMR mutations and germline MMR pathogenic variants, while capturing therapeutic targets, although supporting cost-effectiveness evidence would be needed.

## Data availability statement

The datasets generated during and/or analyzed during the current study are available from the corresponding author on reasonable request.

## Data Availability

The datasets used and/or analyzed during the current study are available from the corresponding author on reasonable request.

## Acknowledgments

We thank members of the Colorectal Oncogenomics Group and members from the Family Cancer Clinics for their support of this manuscript. We thank the participants and staff from the ANGELS and Muir-Torre studies. We thank the Melbourne Bioinformatics and Australian Genome Research Facility for their collaboration on this project. We thank Prof. Ulrike Peters and members of the Genetic Epidemiology of Colorectal cancer Consortium (GECCO) for sharing their targeted multi-gene panel sequencing design that was modified in this study.

