## Supplementary material for "A tumor focused approach to resolving the etiology of DNA mismatch repair deficient tumors classified as suspected Lynch syndrome": Supplementary material_v2.pdf

### METHODS

#### *Pathology Characterization*

For each tumor, pathology reports and Hematoxylin and Eosin (H&E) slides were reviewed by a Pathologist (CR and SD) to determine tumor primary site, tumor grade, histological type, FIGO (International Federation of Gynecology and Obstetrics) stage and sebaceous skin lesion type. In instances where data was missing, tumors were re-staged according to the FIGO 2009 criteria (Pecorelli, 2009).

#### *DNA Mismatch Repair Protein Immunohistochemistry*

DNA mismatch repair (MMR) immunohistochemistry (IHC) testing to categorize the tumor as MMR-deficient (dMMR) as part of the SLS diagnosis was performed by various diagnostic pathology services across Australia and New Zealand prior to study. For the study and where tissue was available, MMR IHC was repeated. Briefly, this testing included staining of 4µm sections on a Ventana *DISCOVERY ULTRA* automated stainer (Ventana Medical Systems Inc., Oro Valley, United States) using anti-MLH1 (M1), anti-MSH2 (G219-1129), anti-MSH6 (SP93) mouse monoclonal and anti-PMS2 (A16-4) rabbit monoclonal primary antibodies (Roche Diagnostics, Basel, Switzerland). All staining protocols were performed according to manufacturer's recommendations (Roche Diagnostics). These MMR IHC results were scored blinded to the original IHC result or tumor panel sequencing results (CR) and tumors were considered dMMR if loss of expression of one or more of the proteins in tumor cell nuclei was observed while lymphocytes and morphologically normal tissue retained protein expression.

*DNA Extraction from Tumor and Blood Biospecimen*

A H&E-stained slide was assessed for each tumor to identify areas of high tumor cellularity for macrodissection. Formalin-fixed paraffin embedded (FFPE) tissue DNA was extracted from the tumor and surrounding normal tissue using the QIAamp DNA FFPE Tissue Kit (Qiagen, Hilden, Germany). DNA was extracted from peripheral blood lymphocytes using the DNeasy blood and tissue kit (Qiagen, Hilden, Germany) and sequenced as a matched germline reference.

*Targeted Multi-Gene Panel Testing*

A multi-gene panel capturing 2.005 megabases (Mb), modified from the capture described in Zaidi *et al.* (Zaidi et al., 2020), was custom-designed for the analysis of tumor and matched blood-derived DNA. The panel comprised of the MMR and *EPCAM* genes as well as other established hereditary CRC and EC genes and consisted of:

Library preparation was performed using the SureSelect$^{XT}$ Low Input Target Enrichment System (Agilent Technologies, Santa Clara, USA) and sequenced on a single lane of a NovaSeq 6000 SP flow cell (Illumina, San Diego, United States) with 300 cycles (150 bp) paired-end reads) at the Australian Genome Research Facility. The median on-target coverage for the 134 test samples was 323.7 for the tumor DNA and 137.4 for blood-derived DNA samples, with an interquartile range of 111.8 – 426.4 and 100.6 – 204.9, respectively. The median on-target coverage for the 53 reference samples was 919.3 (interquartile range 694.6 – 1164.9) for tumor DNA samples and 160.6 (135.8 – 178.0) for blood-derived DNA samples.

*Bioinformatic Analysis*

Adapter sequences were trimmed from raw FASTQ files using Trimmomatic (v.0.38) (Bolger et al., 2014) and aligned to the GRCh37 human reference genome using Burrows-Wheeler Aligner (v.0.7.12) to generate BAM files. Germline and somatic single-nucleotide variants (SNVs) and INDELs were called using Strelka (v.2.9.2, Illumina, San Diego, USA) using the recommended workflow (Saunders et al., 2012). Loss of heterozygosity (LOH) over the MMR, *POLD1* and *POLE* genes were called using the "LOHdeTerminator" (v.0.6, https://github.com/supernifty/LOHdeTerminator) and manually confirmed in BAM files using the Integrative Genomics Viewer (v.2.3). Variants were filtered for PASS with a minimum variant allele fraction of 0.1 and 50x minimum coverage. All variants of interest were verified by manual assessments.

Identified somatic single nucleotide variants (SNVs) and small insertions / deletions (INDELs) were used to calculate TMS according to the simulated annealing approach described by Huang *et al.* (Huang et al., 2018). Single Base Substitutions (TMS SBS) and ID signatures from COSMIC (v.3.2, https://cancer.sanger.ac.uk/signatures/, last accessed date: June 15$^{th}$, 2022) (Tate et al., 2019) were calculated following previous publications (Alexandrov et al., 2013, 2020; Alexandrov & Stratton, 2014).

Tumor mutation burden (TMB) defined as the combined number of somatic SNVs and small insertions / deletions (INDELs) mutations per Mb was calculated as the number of PASS variants as called by Strelka where ≥10 mutations/Mb was considered hypermutated and ≥100 mutations/Mb considered ultra-hypermutated as previously determined (Campbell et al., 2017).

Details of the somatic pipeline and mutational signature calculation are available at https://github.com/supernifty/somatic_pipeline (v.0.3) and https://github.com/supernifty/mutational_signature (v.0.8), respectively. Single Base

Substitutions (SBS) and small (1 to 50 bp) Insertions and Deletions (ID) TMS as described in COSMIC (v.3.2, https://cancer.sanger.ac.uk/signatures/, last accessed date: June 15th, 2022) (Tate et al., 2019) were calculated using the simulated annealing approach described by Huang *et al.* (Huang et al., 2018).

Variants were published in the Variant Call Format according to HGVS nomenclature guidelines (den Dunnen et al., 2016) using canonical RefSeq transcripts GRCh37/hg19 as determined by the Ensembl Variant Effect Predictor (McLaren et al., 2016). Following RefSeq transcripts were used for the candidate genes: *MLH1*: NM_000249.3, *MSH2*: NM_000251.2, *MSH6*: NM_000179.2, *PMS2*: NM_000535.5 and *POLD1*: NM_002691.4. For *BRAF*, *MUTYH* and *POLE* genes, we used following exceptions as these are the more commonly used transcripts: *BRAF:* NM_004333.4, *MUTYH:* NM_001128425.1 and *POLE:* NM_002692.4.

*Determination of DNA Mismatch Repair Status Using the Additive Feature Combination Approach*

Overall tumor dMMR status was determined by applying the additive feature combination approach described in Walker *et al.*, (Walker et al., 2022), where ≥3 features with positivity for dMMR out of the six assessed tools/features, namely MSMuTect (Maruvka et al., 2017), MANTIS (Kautto et al., 2017), MSIseq (Ni Huang et al., 2015), MSISensor (Niu et al., 2014), INDEL count and TMS ID2+ID7 (Georgeson et al., 2021) was considered dMMR. A statistical analysis was performed to determine the optimal thresholds for differentiating dMMR from pMMR tumor status for each tumor type as reported previously (Walker et al., 2022). The thresholds or optimal cut-offs for each of the six tools/features that comprise the additive feature combination approach are shown in the table below (**Supplementary Table S1**).

*Categorization of SLS Cases Using the Accumulated Molecular Results*

The SLS cases were categorized using the results from tumor sequencing as well as re-testing of *MLH1* methylation and MMR IHC as follows:

1) dMMR with a germline pathogenic variant identified (as determined by ClinVar, https://www.ncbi.nlm.nih.gov/clinvar/, last accessed date: December 7th, 2022) (Lynch syndrome, “dMMR-LS”),

2) dMMR with tumor *MLH1* methylation (*MLH1* methylated, “dMMR-MLH1me”),

3) dMMR with tumor and blood *MLH1* methylation (primary epimutation, “dMMR-PriEpi”),

4) dMMR with double somatic MMR variants in the same MMR gene (double somatic mutation, “dMMR-DS”),

5) dMMR with a single somatic MMR variant (single somatic mutation, “dMMR-SS”),

6) dMMR with no germline or somatic variants (suspected Lynch syndrome, “dMMR-SLS”) and

7) pMMR tumors with neither germline or somatic mutations nor hypermethylation of the *MLH1* gene (DNA mismatch repair proficient, “pMMR”).

*PREMM5: Lynch Syndrome Prediction Model*

The PREMM5 scores, clinically used to predict the likelihood for Lynch syndrome in patients of interest, were calculated using the revised model as described by Kastrinos *et al.* (Kastrinos et al., 2017) and implemented using their website (https://premm.dfci.harvard.edu/, last accessed date: 13th November, 2022). A recommended cut-off of ≥2.5% was used to predict Lynch syndrome (Kastrinos et al., 2017).

Weren, R. D. A., Ligtenberg, M. J. L., Kets, C. M., de Voer, R. M., Verwiel, E. T. P., Spruijt, L., van Zelst-Stams, W. A. G., Jongmans, M. C., Gilissen, C., Hehir-Kwa, J. Y., Hoischen, A., Shendure, J., Boyle, E. A., Kamping, E. J., Nagtegaal, I. D., Tops, B. B. J., Nagengast, F. M., Geurts van Kessel, A., van Krieken, J. H. J. M., …

Hoogerbrugge, N. (2015). A germline homozygous mutation in the base-excision repair gene NTHL1 causes adenomatous polyposis and colorectal cancer. *Nature Genetics*, *47*(6), Article 6. https://doi.org/10.1038/ng.3287

Xie, T., D' Ario, G., Lamb, J. R., Martin, E., Wang, K., Tejpar, S., Delorenzi, M., Bosman, F. T., Roth, A. D., Yan, P., Bougel, S., Di Narzo, A. F., Popovici, V., Budinská, E., Mao, M., Weinrich, S. L., Rejto, P. A., & Hodgson, J. G. (2012). A comprehensive characterization of genome-wide copy number aberrations in colorectal cancer reveals novel oncogenes and patterns of alterations. *PloS One*, *7*(7), e42001. https://doi.org/10.1371/journal.pone.0042001

Zaidi, S. H., Harrison, T. A., Phipps, A. I., Steinfelder, R., Trinh, Q. M., Qu, C., Banbury, B. L., Georgeson, P., Grasso, C. S., Giannakis, M., Adams, J. B., Alwers, E., Amitay, E. L., Barfield, R. T., Berndt, S. I., Borozan, I., Brenner, H., Brezina, S., Buchanan, D. D., … Peters, U. (2020). Landscape of somatic single nucleotide variants and indels in colorectal cancer and impact on survival. *Nature Communications*, *11*(1), 3644. https://doi.org/10.1038/s41467-020-17386-z

### Table Legends

**Supplementary Table S1.** Table displaying optimal cut-offs for the six tumor features determined previously (Walker et al., 2022) in the additive feature combination approach. Abbreviations: colorectal cancer, CRC; endometrial cancer, EC; sebaceous skin tumor, SST; DNA mismatch repair deficient, dMMR; DNA mismatch repair proficient, pMMR; insertions / deletions, INDELs; tumor mutational signatures, TMS; small (1 to 50 base pair) insertions / deletions, IDs; single base substitution, SBS and tumor mutational burden, TMB.

| | CRC | EC | SST |
|---|---|---|---|
| Tumor Features | pMMR < n ≥ dMMR | pMMR < n ≥ dMMR | pMMR < n ≥ dMMR |
| MSMuTect | 48 | 31 | 25 |
| MSIseq | 2.494 | 0.499 | 0.499 |
| MANTIS | 0.252 | 0.274 | 0.217 |
| INDEL count | 5 | 1 | 3 |
| MSISensor | 6.9 | 21.21 | 2.05 |
| TMS ID2+ID7 | 0.504 | 0.182 | 0.547 |

**Supplementary Table S2.** SLS tumors (n=13) that showed discordant MMR IHC findings between clinical diagnostic testing before study entry and testing completed internally during this study and the change in their MMR status and/or pattern of MMR protein loss Abbreviations: colorectal cancer, CRC; endometrial cancer, EC; sebaceous skin tumor, SST; DNA mismatch repair, MMR; DNA mismatch repair deficient, dMMR; DNA mismatch repair proficient, pMMR; not tested, NT. + Indicates heterogeneous loss of MMR protein expression by IHC.

| | | | **Pre-study Clinical MMR IHC and *MLH1* Methylation Testing Results** | | **Internally Performed MMR IHC and *MLH1* Methylation Testing Results** | | | **Results from Tumor Panel Sequencing Performed during the Study** | |
|---|---|---|---|---|---|---|---|---|---|
| **No.** | **ID** | **Tumor type** | **IHC Pattern** | ***MLH1* Methylation (MS-MLPA)[a]** | **IHC Pattern** | ***MLH1* *M*ethylation (MethyLight [PMR])** | ***MLH1* Methylation (MS-HRM [%])** | **dMMR Status from Additive Tumor Feature Approach** | **Cause for DNA Mismatch Repair Deficiency** |
| 1 | ID001 | CRC | MLH1 / PMS2 | NEGATIVE | Normal | NEGATIVE (0) | NEGATIVE (0) | pMMR (0/6) | - |
| 2 | ID002 | CRC | MSH2 / MSH6 | NT | Normal | NT | NT | pMMR (2/6) | - |
| 3 | ID003 | CRC | MSH2 / MSH6 | NT | Normal | NT | NT | pMMR (0/6) | - |
| 4 | ID004 | CRC | MSH6 | NT | Normal | NT | NT | pMMR (0/6) | - |

| | | | | | | | | | |
|---|---|---|---|---|---|---|---|---|---|
| 5 | ID005 | CRC | MSH6 | NT | Normal MSH6 expression | NT | NT | pMMR (1/6) | - |
| 6 | ID006 | EC | MSH6 | NT | Normal | NT | NT | pMMR (0/6) | - |
| 7 | ID007 | CRC | PMS2 | NT | Normal | NEGATIVE (0) | NEGATIVE (0) | pMMR (0/6) | - |
| 8 | ID008 | CRC | MSH6 and PMS2 | NT | Normal | NT | NT | pMMR (0/6) | - |
| 9 | ID009 | CRC | MSH6 | NT | MLH1 / PMS2 | POSITIVE (79) | POSITIVE (40) | dMMR (6/6) | *MLH1* methylated tumor. |
| 10 | ID010 | CRC | PMS2 | NT | MLH1 / PMS2 | NEGATIVE (0) | NEGATIVE (0) | dMMR (6/6) | Double somatic hit in *MLH1*. |
| 11 | ID011 | EC | MSH6 | NT | $MLH1^{+}$ / PMS2 and MSH6 | POSITIVE (26) | POSITIVE (5) | pMMR (2/6) | This person carries a germline pathogenic *MSH6* variant with a somatic *MSH6* mutation (2nd hit) and is positive for tumor *MLH1* methylation. |

| | | | | | | | | | |
|---|---|---|---|---|---|---|---|---|---|
| 12 | ID012 | CRC | MSH6 and PMS2 | NT | MSH6[+] | NT | NT | pMMR (1/6) | Double somatic hit in *MSH6*. |
| 13 | ID013 | SST | PMS2 | NT | MLH1[b] | NEGATIVE (0) | NEGATIVE (0) | dMMR (6/6) | Double somatic hit in *MLH1*. |

*[+] Indicates heterogeneous loss of MMR protein expression by IHC.*

*[a] All pre-study clinical MLH1 methylation results were generated using methylation specific - multiplexed ligation dependent probe amplification assay (MS-MLPA), unless where indicated with BRAF V600E testing.*

*[b] Only MLH1 protein expression was tested due to only a single slide available for IHC.*

**Supplementary Table S3.** The concordance between the final MMR IHC result and the predicted dMMR status from the additive feature combination approach overall and by tumor type. Abbreviations: colorectal cancer, CRC; endometrial cancer, EC; sebaceous skin tumor, SST; DNA mismatch repair deficient, dMMR; DNA mismatch repair proficient, pMMR.

| | **Additive Feature Combination Approach for dMMR Prediction** | | | | | | | |
|---|---|---|---|---|---|---|---|---|
| | **Tumor type combined** | **Total** | **CRC n=80** | **Total CRC** | **EC n=33** | **Total EC** | **SST n=24** | **Total SST** |

| **Final MMR IHC status** | *dMMR* | *pMMR* | | *dMMR* | *pMMR* | | *dMMR* | *pMMR* | | *dMMR* | *pMMR* | |
|---|---|---|---|---|---|---|---|---|---|---|---|---|
| *MLH1/PMS2 loss* | 58 | 7 | ***65*** | 37 | 1 | **38** | 13 | 6 | **19** | 8 | 0 | **8** |
| *MLH1/PMS2 and MSH6 loss* | 0 | 1 | ***1*** | 0 | 0 | **0** | 0 | 1 | **1** | 0 | 0 | **0** |
| *MSH2/MSH6 loss* | 47 | 0 | ***47*** | 27 | 0 | **27** | 5 | 0 | **5** | 15 | 0 | **15** |
| *MSH6 only loss* | 9 | 3 | ***12*** | 3 | 1 | **4** | 5 | 2 | **7** | 1 | 0 | **1** |
| *PMS2 only loss* | 1 | 1 | ***2*** | 1 | 1 | **2** | 0 | 0 | **0** | 0 | 0 | **0** |
| *All four protein loss* | 2 | 0 | ***2*** | 2 | 0 | **2** | 0 | 0 | **0** | 0 | 0 | **0** |
| *pMMR* | 0 | 8 | ***8*** | 0 | 7 | **7** | 0 | 1 | **1** | 0 | 0 | **0** |
| **Total** | **117** | **20** | **137** | **70** | **10** | **80** | **23** | **10** | **33** | **24** | **0** | **24** |

**Supplementary Table S4.** The tumor *MLH1* methylation testing completed for SLS tumors prior to entering the study showing either negative, inconclusive, or not tested results and the subsequent MLH1 methylation testing results from internal testing using MethyLight and MS-HRM assays highlighting the positive MLH1 methylation results found by this study. Abbreviations: colorectal cancer, CRC; endometrial cancer, EC; sebaceous skin tumor, SST; DNA mismatch repair, MMR; DNA mismatch repair deficient, dMMR; DNA mismatch repair proficient, pMMR; BRAF V600E, BRAF; not tested, NT; not applicable, NA. + Indicates heterogeneous loss of MMR protein expression by IHC.

| | | | | | | Pre-study Clinical MMR IHC and *MLH1* Methylation Testing Results | | Internally performed *MLH1* Methylation and MMR IHC | | | Results from Tumor Panel Sequencing Performed During the Study | | |
|---|---|---|---|---|---|---|---|---|---|---|---|---|---|
| | | No. | ID | Tumor type | Age at Diagnosis | IHC Pattern | *MLH1* Methylation (MS-MLPA)[a] | IHC Pattern | *MLH1* Methylation (MethyLight [PMR-value ≥10%]) | *MLH1* Methylation (MS-HRM [%-value ≥5%]) | dMMR Status (Additive Tumor Feature Approach) | *BRAF* V600E Status | Cause for Tumour DNA Mismatch Repair Deficiency |
| Clinical | **negative** | 1 | ID014 | EC | 66-70 | MLH1/ PMS2 | **NEGATIVE** | NT | **POSITIVE (34)[c]** | **POSITIVE (5)[c]** | dMMR (5/6) | NA | *MLH1* methylated tumor. |

| | | | | | | | | | | | | |
|---|---|---|---|---|---|---|---|---|---|---|---|---|
| | | 2 | ID015 | EC | 71-75 | MLH1/ PMS2 | **NEGATIVE** | NT | **POSITIVE (65)**[c] | **POSITIVE (20)**[c] | dMMR (6/6) | NA | *MLH1* methylated tumor. |
| | | 3 | ID016 | EC | 71-75 | MLH1/ PMS2 | **NEGATIVE** | NT | **POSITIVE (19)**[c] | **POSITIVE (10)**[c] | pMMR (1/6) | NA | *MLH1* methylated tumor. |
| | | 4 | ID017 | EC | 76-80 | MLH1/ PMS2 | **NEGATIVE** | MLH1/ PMS2 | **POSITIVE (17)**[c] | **POSITIVE (10)**[c] | pMMR (2/6) | NA | *MLH1* methylated tumor. |
| | | 5 | ID018 | CRC | 21-25 | Loss of all four MMR protein s | **NEGATIVE** | NT | **POSITIVE (34)**[e] | **POSITIVE (20)**[e] | dMMR (6/6) | Wildtype | Carrier of a germline pathogenic variant in MSH2 with somatic MSH2 mutation (2nd hit), where CRC tumor was also positive for MLH1 methylation. |
| Clinical | **inconclusive** | 6 | ID019 | CRC | 41-45 | MLH1/ PMS2 | **INCONCLU SIVE**[b] | NT | **POSITIVE (24)**[e] | **POSITIVE (5)**[e] | dMMR (6/6) | Wildtype | *MLH1* methylated tumor. |

| | | | | | | | | | | | | | |
|---|---|---|---|---|---|---|---|---|---|---|---|---|---|
| | | 7 | ID020 | CRC | 41-45 | MLH1/ PMS2 | **INCONCLUSIVE** | NT | **POSITIVE (19)**[c] | **POSITIVE (30)**[c] | dMMR (6/6) | Wildtype | *MLH1* methylated tumor. |
| | | 8 | ID021 | EC | 71-75 | MLH1/ PMS2 | **INCONCLUSIVE** | MLH1/ PMS2 | **POSITIVE (78)**[c] | **POSITIVE (10)**[c] | dMMR (6/6) | NA | *MLH1* methylated tumor. |
| | | 9 | ID022 | EC | 51-55 | MLH1/ PMS2 | **INCONCLUSIVE**[f] | MLH1/ PMS2 | **POSITIVE (107)**[c] | **POSITIVE (10)**[c] | dMMR (3/6) | NA | *MLH1* methylated tumor. |
| | | 10 | ID023 | EC | 51-55 | MLH1/ PMS2 | **INCONCLUSIVE** | NT | **POSITIVE (55)**[c] | **POSITIVE (20)**[c] | dMMR (4/6) | NA | *MLH1* methylated tumor. |
| | | 11 | ID024 | EC | 71-75 | MLH1/ PMS2 | **INCONCLUSIVE** | NT | **POSITIVE (10)**[c] | **POSITIVE (5)**[c] | pMMR (1/6) | NA | *MLH1* methylated tumor. |
| | | 12 | ID025 | CRC | 36-40 | MLH1/ PMS2 | **INCONCLUSIVE** | MLH1/ PMS2 | **NEGATIVE (0)**[c] | **NEGATIVE (0)**[c] | dMMR (6/6) | Wildtype | Double somatic mutations in *MLH1*. |
| Clinical | **not** | 13 | ID026 | CRC | 56-60 | MLH1/ PMS2 | **NEGATIVE**[g] | NT | **POSITIVE (32)**[c] | **POSITIVE (5)**[c] | dMMR (4/6) | Wildtype | *MLH1* methylated tumor. |

| | | | | | | | | | | | | | |
|---|---|---|---|---|---|---|---|---|---|---|---|---|---|
| | | 14 | ID027 | CRC | 36-40 | MLH1/ PMS2 | **NT (*BRAF* NEGATIVE)** | NT | **POSITIVE (15)**[e] | **POSITIVE (20)**[e] | dMMR (5/6) | Wildtype | *MLH1* methylated tumor. |
| | | 15 | ID028 | CRC | 51-55 | MLH1/ PMS2 | **NT (*BRAF* INCONCLUSIVE)** | NT | **POSITIVE (36)**[c] | **POSITIVE (30)**[c] | dMMR (6/6) | V600E | *MLH1* methylated tumor. |
| | | 16 | ID029 | CRC | 51-55 | MLH1/ PMS2 | **NT** | NT | **POSITIVE (74)**[c] | **POSITIVE (30)**[c] | dMMR (6/6) | Wildtype | *MLH1* methylated tumor. |
| | | 17 | ID030 | CRC | 31-35 | MLH1/ PMS2 | **NT** | NT | **POSITIVE (66)**[e] | **POSITIVE (30)**[e] | dMMR (6/6) | Wildtype | *MLH1* methylated tumor. |
| | | 18 | ID009 | CRC | 61-65 | MSH6 | **NT** | MLH1/ PMS2 | **POSITIVE (79)**[c] | **POSITIVE (40)**[c] | dMMR (6/6) | V600E | *MLH1* methylated tumor. |
| | | 19 | ID031 | CRC | 31-35 | PMS2 | **NT** | PMS2 | **POSITIVE (34)**[e] | **POSITIVE (5)**[e] | pMMR (1/6) | V600E | *MLH1* methylated tumor. |
| | | 20 | ID032 | EC | 56-60 | MLH1/ PMS2 | **NT** | MLH1/ PMS2 | **POSITIVE (83)**[c] | **POSITIVE (30)**[c] | dMMR (6/6) | NA | *MLH1* methylated tumor. |

| | | | | | | | | | | | | | |
|---|---|---|---|---|---|---|---|---|---|---|---|---|---|
| | | 21 | ID011 | EC | 56-60 | MSH6 | **NT** | MLH1$^{+}$/ PMS2 and MSH6 | **POSITIVE (26)**[c] | **POSITIVE (5)**[c] | pMMR (2/6) | NA | Carrier of a germline pathogenic variant in MSH6 with somatic MSH6 mutation (2nd hit) and was positive for tumor MLH1 methylation. |
| | | 22 | ID033 | SST | 76-80 | MLH1/ PMS2 | **NT** | NT | **SST: POSITIVE (57) Blood: POSITIVE (3)**[d] | **SST: POSITIVE (40) Blood: POSITIVE (10)** | dMMR (6/6) | NA | *Primary epimutation* - SST and blood DNA both *MLH1* methylation positive |
| | | 23 | ID013 | SST | 61-65 | MLH1/ PMS2 | **NT** | NT | **SST: POSITIVE (19) Blood:** | **SST: POSITIVE (20) Blood:** | dMMR (6/6) | NA | *Primary epimutation* – SST, CRC and blood |

| | | | | | | | | | | | | | |
|---|---|---|---|---|---|---|---|---|---|---|---|---|---|
| | | | | | | | | | **POSITIVE (32)** | **POSITIVE (30)** | | | DNA *MLH1* methylation positive |
| | | | | CRC | 51-55 | MLH1/ PMS2 | **NT** | NT | **CRC: POSITIVE (75)** | **CRC: POSITIVE (40)** | dMMR (6/6) | Wildtype | |

*[+] Indicates heterogeneous loss of MMR protein expression by IHC.*

*[a] All clinical MLH1 methylation results were generated using methylation specific - multiplexed ligation dependent probe amplification assay (MS-MLPA), unless where indicated that BRAF V600E testing was used as a surrogate for MLH1 methylation testing.*

*[b] Clinics indicated DNA quality as low.*

*[c] Blood DNA was not tested for MLH1 methylation.*

*[d] In-house testing showed low levels of MLH1 methylation but called positive overall.*

*[e] Blood-derived DNA was tested for MLH1 methylation and found to be negative for MLH1 methylation.*

*[f] Repeated twice.*

*[g] Only tested on blood-derived DNA, no tumor testing was performed.*

**Supplementary Table S5.** Presentation of germline pathogenic variants and variants of uncertain clinical significance (VUS) identified in the MMR, *MUTYH* and *POLE* genes. Abbreviations: colorectal cancer, CRC; endometrial cancer, EC; sebaceous skin tumor, SST; DNA mismatch repair, MMR; DNA mismatch repair deficient, dMMR; DNA mismatch repair proficient, pMMR; loss of heterozygosity, LOH; tumor mutational signature, TMS; variant of uncertain clinical significance, VUS; Lynch syndrome, LS; *MLH1* methylation, dMMR-MLH1me; double somatic mutation, dMMR-DS.

| No. | ID | Tumor Type | Sex | AgeDx | **FINAL MMR IHC Status** | MMR Status by Additive Feature Approach | **Germline Variant** | VarSome ACMG Implementation for Germline Variant | New Proposed Variant Classification | Comments | **FINAL Tumor / Individual Categorization** |
|---|---|---|---|---|---|---|---|---|---|---|---|
| 1 | ID018 | CRC | M | 21-25 | **Loss of all four MMR proteins** | dMMR (3/6) | ***MSH2*: deletion of exon 7** | - | PATHOGENIC | Somatic mutation (2nd hit) *MSH2*: c.2458+1G>A p.? | **dMMR - LS + (dMMR - MLH1me)** |

| | | | | | | | | | | | |
|---|---|---|---|---|---|---|---|---|---|---|---|
| 2 | ID034 | EC | F | 56-60 | **MLH1/PMS2 and MSH6** | pMMR (2/6) | ***MSH6*: c.3834_3849dup p.Thr1284Glnfs*10** | Pathogenic (Class 5): PVS1 \| PM2 \| PP3 | PATHOGENIC | Somatic mutation (2nd hit) *MSH6*: c.3261del p.Phe1088Serfs*2 | **dMMR - LS + (dMMR - MLH1me)** |
| 3 | ID013 | SST | M | 71-75 | **MLH1/PMS2** | dMMR (6/6) | ***MLH1*: c.400A>G p.Lys134Glu** | Uncertain Significance (Class 3): PM1 \| PP3 \| PM2 | VUS | LOH across *MLH1* \| somatic mutation *MLH1*: c.199G>A p.Gly67Arg | **dMMR - DS (MLH1)** |
| 4 | ID035 | CRC | F | 41-45 | **MSH2/MSH6** | dMMR (6/6) | ***MLH1*: c.1153C>T p.Arg385Cys** | Uncertain Significance (Class 3): PP5 \| PM2 \| PP2 \| PP3 | VUS | No somatic *MLH1* mutations | **dMMR - DS (MSH2)** |

| | | | | | | | | | | | |
|---|---|---|---|---|---|---|---|---|---|---|---|
| 5 | ID036 | CRC | F | 36-40 | **MLH1/PMS2** | dMMR (6/6) | ***MSH2*: c.668T>C p.Leu223Pro** | Uncertain Significance (Class 3): PP3 \| PM2 | VUS | No somatic *MSH2* mutations | **dMMR - DS (MLH1)** |
| 6 | ID037 | CRC | F | 41-45 | **MLH1/PMS2** | dMMR (6/6) | ***MSH6*: c.2827G>T p.Asp943Tyr** | Uncertain Significance (Class 3): PM2 \| PP3 | VUS | No somatic *MSH6* mutations | **dMMR – DS (MLH1)** |
| 7 | ID027 | CRC | F | 36-40 | **MLH1/PMS2** | dMMR (5/6) | ***MSH6*: c.2963G>A p.Arg988His** | Uncertain Significance (Class 3): PM2 \| BP4 | VUS | Somatic mutation in *MSH6*: c.2963G>A p.Arg988His | **dMMR - MLH1me** |
| 8 | ID038 | EC | F | 61-65 | **MLH1/PMS2** | dMMR (5/6) | ***MSH6*: c.*85T>A p.?** | Uncertain significance (Class 3): PM2 \| BP4 | VUS | Somatic mutation in *MSH6*: c.3261del | **dMMR - DS (MLH1)** |

| | | | | | | | | | | | |
|---|---|---|---|---|---|---|---|---|---|---|---|
| | | | | | | | | | | p.Phe1088Serfs*2 | |
| 9 | ID039 | SST | M | 66-70 | **MSH2/MSH6** | dMMR (6/6) | ***PMS2*: c.*3G>A p.?** | Uncertain Significance (Class 3): PM2 \| BP4 | VUS | No somatic *PMS2* mutations | **dMMR - DS (MSH2)** |
| 10 | ID040 | SST | M | 71-75 | **MSH2/MSH6** | dMMR (5/6) | ***MSH2*: c.138C>G p.His46Gln** | Uncertain Significance (Class 3): PS1 \| PM2 \| PP2 \| PP3 \| PP5 \| BP6 | LIKELY BENIGN | LOH across *MSH2* \| MSH2: c.1759G>C p.Gly587Arg | **dMMR - DS (MSH2)** |
| 11 | ID041 | EC | F | 61-65 | **MSH6** | dMMR (3/6) | ***MSH2*: c.138C>G p.His46Gln** | Uncertain Significance (Class 3): PS1 \| PM2 \| PP2 \| PP3 \| PP5 \| BP6 | LIKELY BENIGN | Somatic mutation in *MSH2*: c.1354G>A p.Glu452Lys | **dMMR - DS (MSH6)** |

| | | | | | | | | | | | |
|---|---|---|---|---|---|---|---|---|---|---|---|
| 12 | ID041 | EC | F | 61-65 | **MSH6** | dMMR (3/6) | ***MUTYH*: c.536A>G p.Tyr179Cys** | Pathogenic (Class 5): PP5 \| PS3 \| PM2 | PATHOGENIC | No *MUTYH* deficiency TMS (SBS18 / SBS36) | **dMMR - DS (MSH6)** |
| 13 | ID042 | EC | F | 61-65 | **MSH2/MSH6** | dMMR (5/6) | ***MUTYH*: c.1187G>A p.Gly396Asp** | Pathogenic (Class 5): PS3 \| PP3 \| PP5 \| PM5 | PATHOGENIC | No *MUTYH* deficiency TMS (SBS18 / SBS36) | **dMMR - DS (MSH2)** |
| 14 | ID043 | SST | M | 71-75 | **MSH2/MSH6** | dMMR (6/6) | ***MUTYH*: c.1187G>A p.Gly396Asp** | Pathogenic (Class 5): PS3 \| PP3 \| PP5 \| PM5 | PATHOGENIC | No *MUTYH* deficiency TMS (SBS18 / SBS36) | **dMMR - DS (MSH2)** |
| 15 | ID001 | CRC | M | 71-75 | **Normal** | pMMR (0/6) | ***MUTYH*: c.1276C>T p.Arg426Cys** | Likely Benign (Class 2): BP4 \| PM2 | VUS | No *MUTYH* deficiency TMS (SBS18 / SBS36) | **pMMR** |

| | | | | | | | | | | | |
|---|---|---|---|---|---|---|---|---|---|---|---|
| 16 | ID044 | CRC | M | 31-35 | **PMS2** | dMMR (6/6) | ***POLE:*** **c.825C>G p.Asp275Glu** | Uncertain Significance (Class 3): PM2 \| PP3 \| BP1 | VUS | No *POLE/POLD1* related TMS; Somatic mutation *POLD1*: c.1327C>T p.Arg443Trp; ultra-hypermutated TMB (437 mutations/Mb) | **dMMR - DS (PMS2)** |
| 17 | ID045 | CRC | M | 56-60 | **MSH2/MSH6** | dMMR (6/6) | ***POLE*****: c.861T>A p.Asp287Glu** | Uncertain Significance (Class 3): PM2 \| PP3\| BP1 | VUS | No *POLE/POLD1* related TMS; somatic *POLD1* mutation c.1054C>T | **dMMR - DS (MSH2)** |

| | | | | | | | | | | |
|---|---|---|---|---|---|---|---|---|---|---|
| | | | | | | | | | | p.Arg352Cys; ultra-hypermutated TMB (118 mutations/Mb) |

**Supplementary Table S6.** Summary of the clinicopathological features for the double somatic MMR mutation (dMMR-DS) tumors overall and by tumor type. Abbreviations: Standard deviation, SD; colorectal cancer, CRC; endometrial cancer, EC; sebaceous skin tumor, SST; DNA mismatch repair, MMR; DNA mismatch repair deficient, dMMR; DNA mismatch repair deficient tumor with double somatic mutations, dMMR-DS.

| **Double Somatic MMR Mutations (dMMR-DS)** | **CRC** | **EC** | **SST** | **Total** |
|---|---|---|---|---|
| Total No. | 56 | 15 | 17 | 88 |
| **Sex, n (%)** | | | | |
| Male | 28 (50%) | 0 (0%) | 16 (94.1%) | 44 (50%) |
| Female | 28 (50%) | 15 (100%) | 1 (5.9%) | 44 (50%) |
| **Age at Tumor Diagnosis, n (%)** | | | | |
| Mean ± SD | 46.6 ± 13.1 | 58.3 ± 7.4 | 65.9 ± 11.6 | 52.3 ± 14.3 |
| Range | 26-74 | 43-72 | 81 | 26-81 |
| Proportion diagnosed ≤ 50 years | 35 (62.5%) | 2 (13.3%) | 3 (17.6%) | 37 (45.5%) |
| **PREMM5 Score, n (%)** | | | | |
| Mean ± SD | 7.2 ± 7.9 | 3.2 ± 1.6 | 2.1 ± 1.4 | 5.6 ± 6.7 |
| Range | 1.1-50 | 1.8-7.2 | 0.8-6.6 | 0.8-50 |
| Proportion ≥2.5 threshold | 48 (85.7%) | 10 (66.7%) | 3 (17.6%) | 61 (69.3%) |
| Proportion ≥5 threshold | 27 (48.2%) | 2 (13.3%) | 1 (5.9%) | 30 (34.1%) |
| **CRC Tumor Site, n (%)** | | | | |
| Proximal | 38 (67.9%) | - | - | - |
| Distal | 13 (23.2%) | - | - | - |
| Rectum | 5 (8.9%) | - | - | - |

| | | | | |
|---|---|---|---|---|
| **SST Site, n (%)** | | | | |
| Head/Neck | - | - | 10 (58.8%) | - |
| Trunk/Limb | - | - | 7 (41.2%) | - |

### Figure Legends

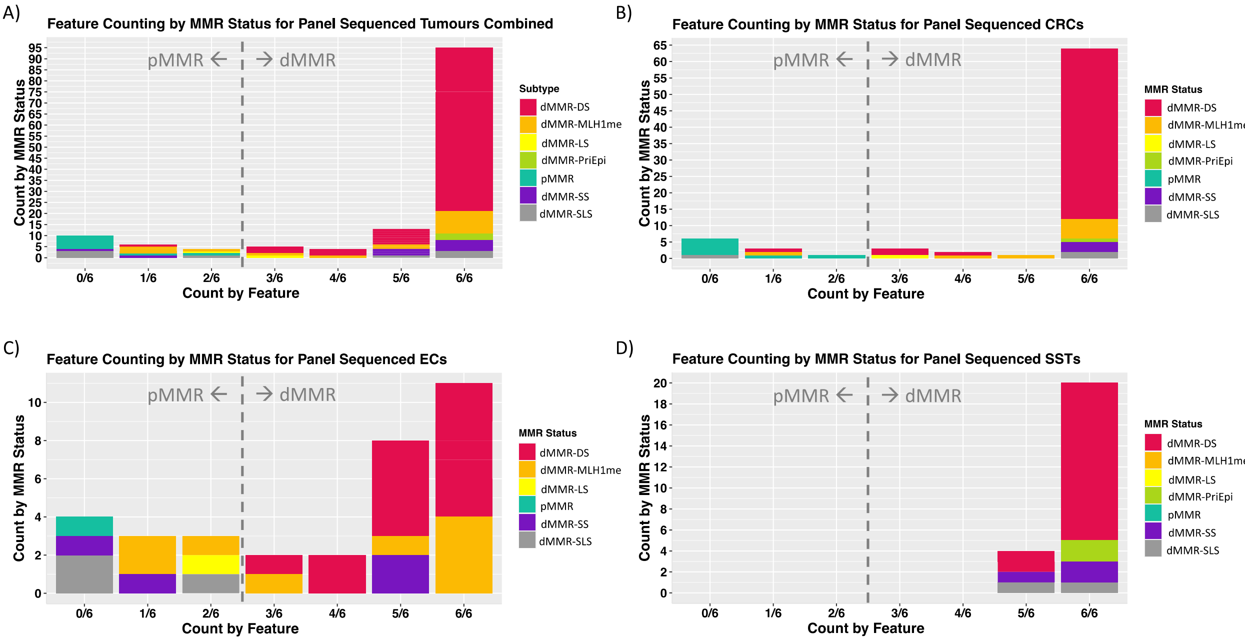

**Supplementary Figure S1:** Bar plots presenting the results from the additive tumor feature combination approach to assess the MMR status in the double somatic mutation cohort for A) all tumors combined and separated by B) CRC, C) EC and D) SST tissue types. Abbreviations: DNA mismatch repair, MMR; colorectal cancer, CRC; endometrial cancer, EC; sebaceous skin tumor, SST; double somatic mutation, dMMR-DS; positive *MLH1* methylation, dMMR-MLH1me; Lynch syndrome, dMMR-LS; primary epimutation, dMMR-PriEpi; DNA mismatch repair proficient, pMMR; single somatic mutation, dMMR-SS; suspected Lynch syndrome, dMMR-SLS.

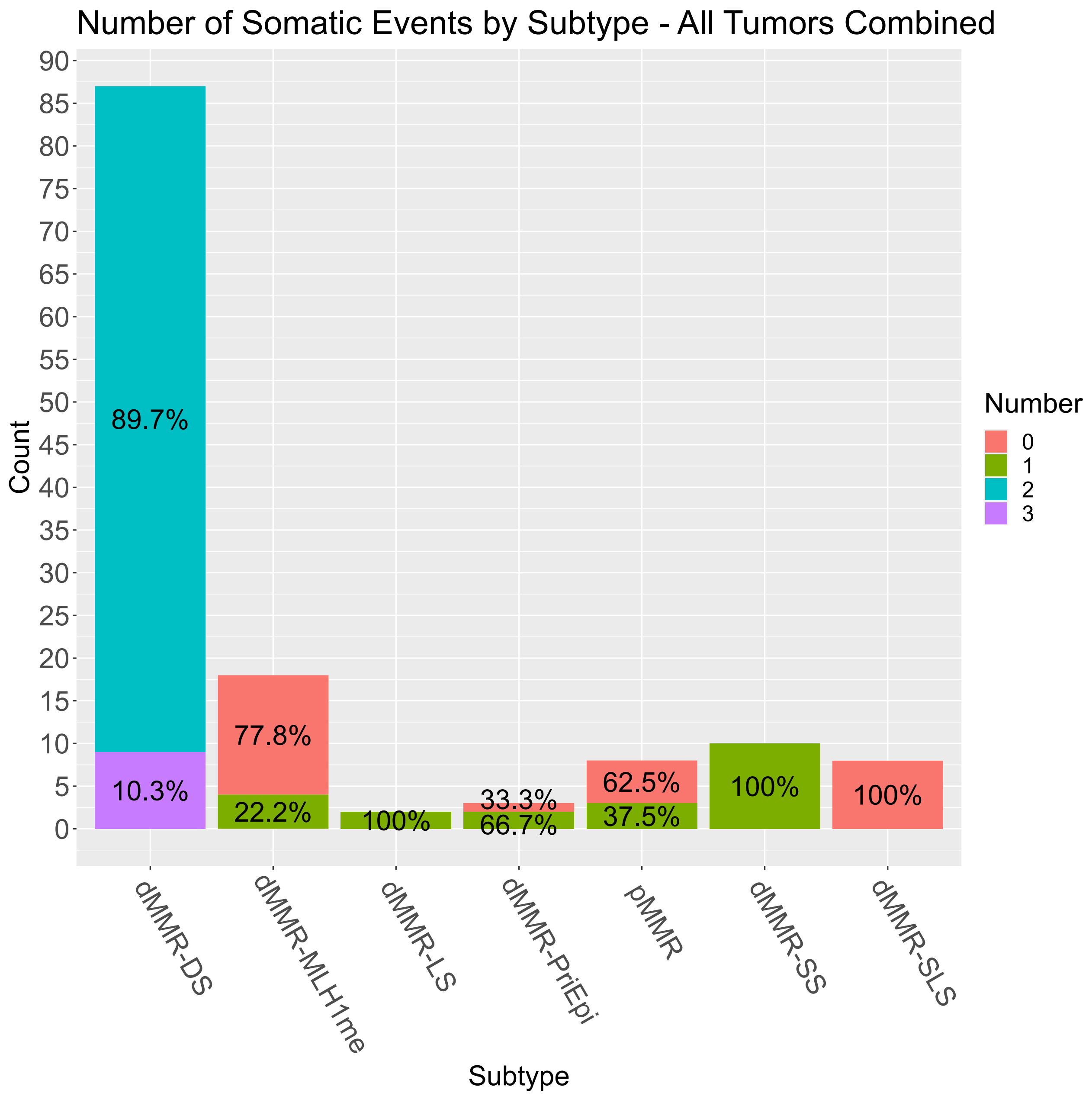

**Supplementary Figure S2:** Bar plot presenting the prevalence of pathogenic / likely pathogenic somatic mutations (including loss of heterozygosity, LOH) by subtype for the study cohort. Excluded tumors that presented with loss of all four MMR proteins (n=2) and MLH1/PMS2 with concurrent MSH6 loss (n=2) by MMR IHC. Number of somatic events were counted fitting to observed loss in MMR IHC. For pMMR, somatic events observed in all four MMR genes were added to somatic variant count. Excluded tumors that presented with loss of all four MMR proteins (n = 2) and MLH1/PMS2 with concurrent MSH6 loss (n = 1) by

MMR IHC. Number of somatic events were counted fitting to observed loss in MMR IHC. For pMMRs, somatic events in all four MMR variants were added to the somatic variant count. Abbreviations: DNA mismatch repair, MMR; immunohistochemistry, IHC.

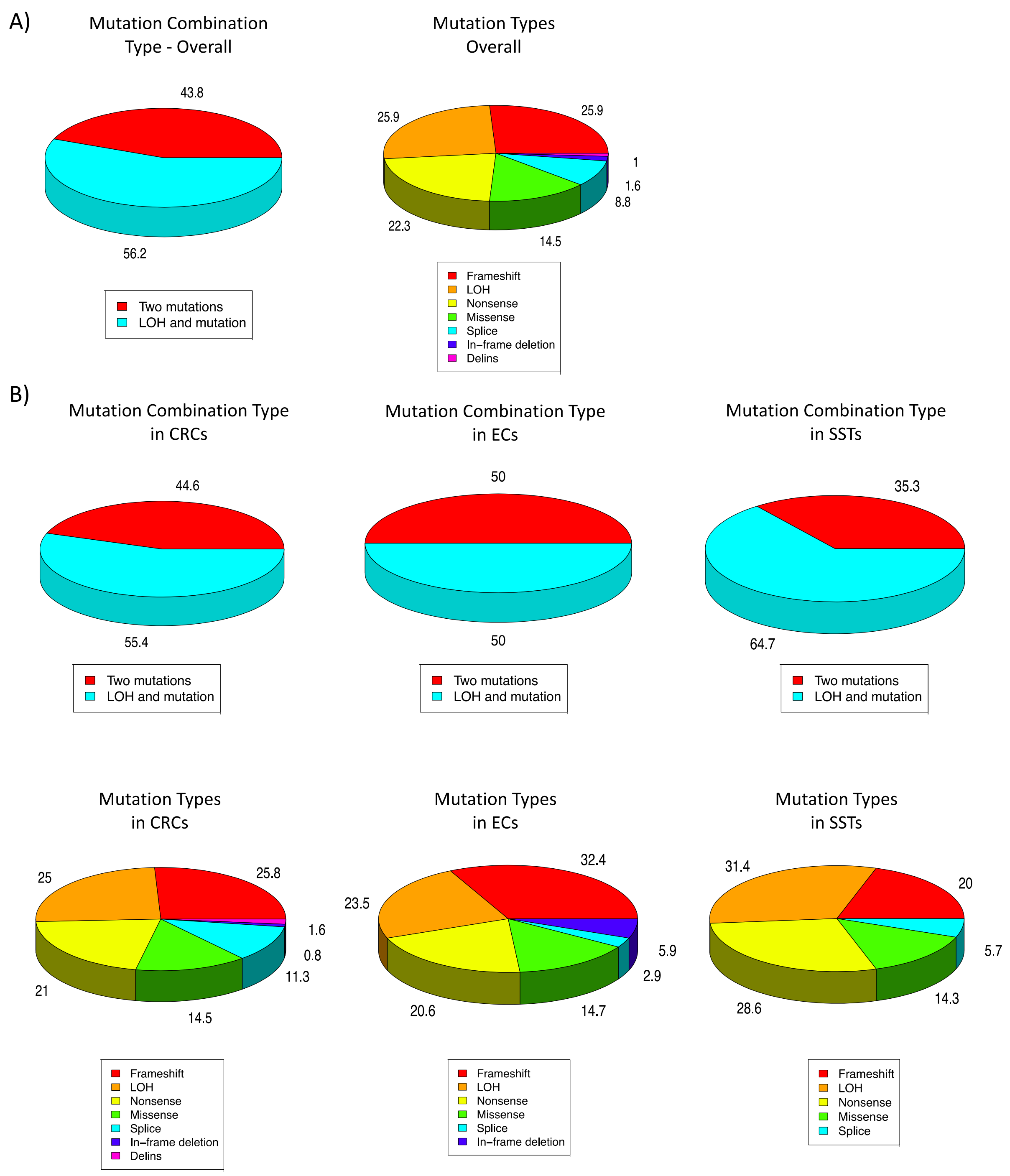

**Supplementary Figure S3:** Pie graphs displaying the frequency of the mutation combination type (two single somatic mutations versus a single somatic mutation with loss of heterozygosity (LOH)) as well as the type of mutation A) overall and B) separated by tissue type. Abbreviations: Loss of heterozygosity, LOH; colorectal cancer, CRC; endometrial cancer, EC; sebaceous skin tumor, SST.

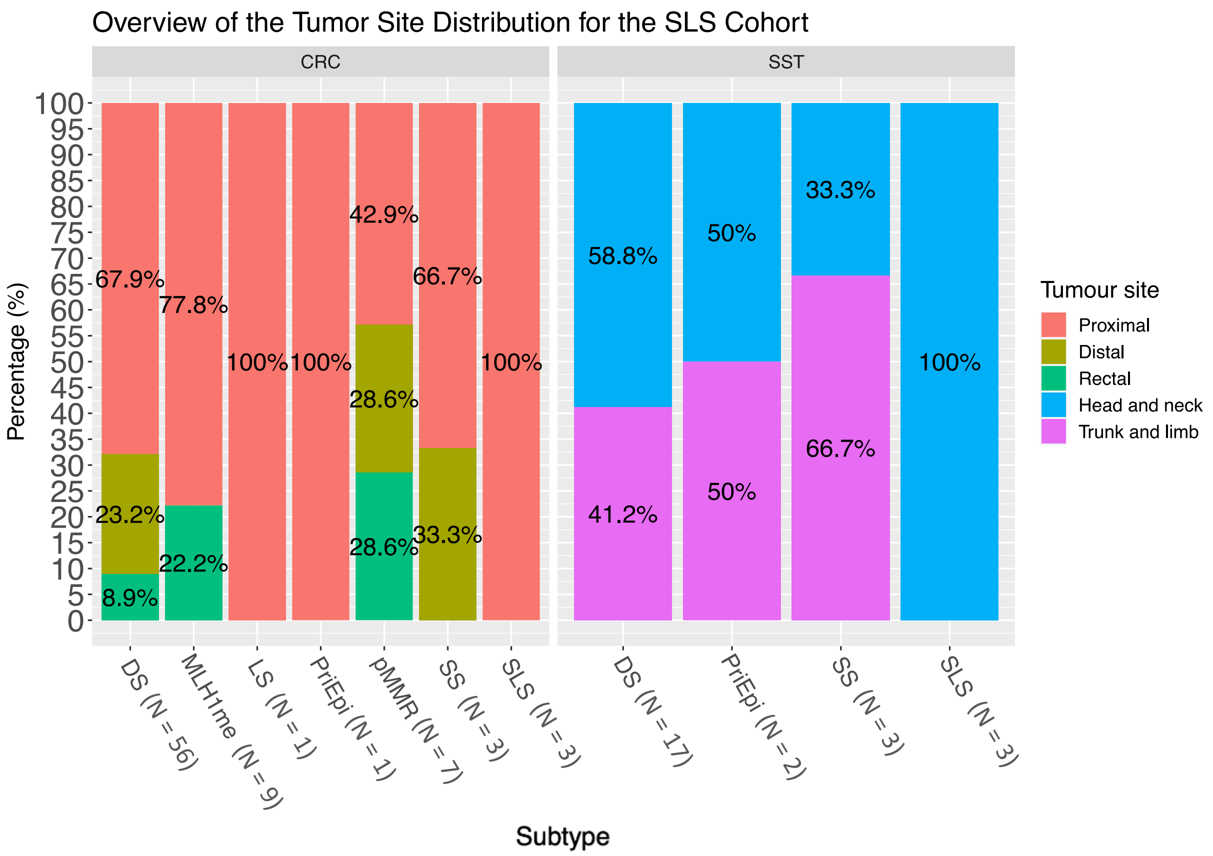

**Supplementary Figure S4:** Bar graphs presenting the site distribution in the double somatic mutation cohort across all CRCs and SSTs. Abbreviations: Colorectal cancer, CRC; sebaceous skin tumor, SST; double somatic mutations, dMMR-DS; positive *MLH1* methylation, dMMR-MLH1me; Lynch syndrome, dMMR-LS; primary *MLH1* epimutation, dMMR-PriEpi; DNA mismatch repair proficient, pMMR; single somatic mutation, dMMR-SS; suspected Lynch syndrome, dMMR-SLS.

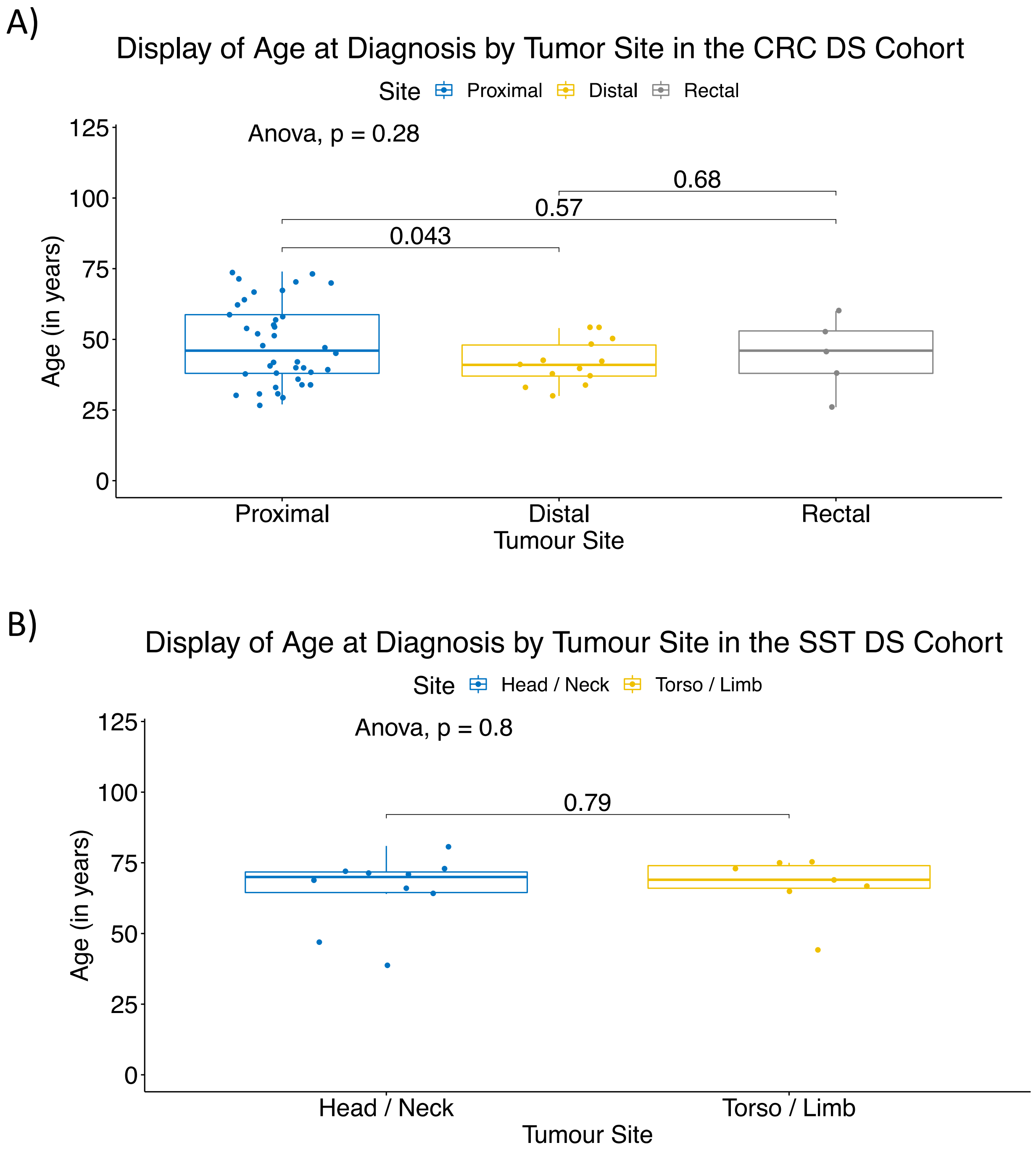

**Supplementary Figure S5:** Boxplots presenting the site distribution in the double somatic mutation cohort across all A) CRCs and B) SSTs. Significant (< 0.05) p-values are indicated for pairwise (t-test) and multigroup comparisons (Anova). Abbreviations: Colorectal cancer, CRC; sebaceous skin tumor, SST; double somatic mutations, DS.

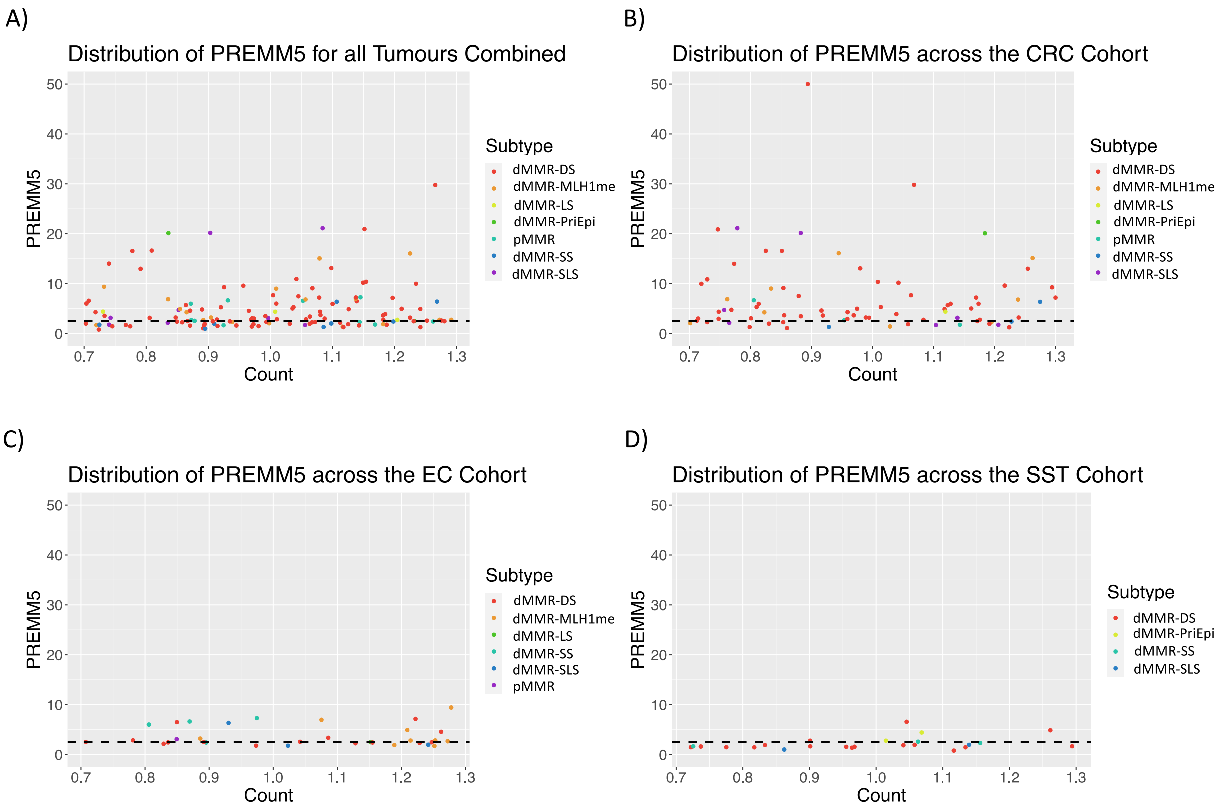

**Supplementary Figure S6:** Scatter plots presenting the PREMM5 score distribution in the test cohort for A) all tumors combined and separated by B) CRC, C) EC and D) SST tissue types. Abbreviations: Colorectal cancer, CRC; endometrial cancer, EC; sebaceous skin tumor, SST; double somatic mutations, dMMR-DS; positive *MLH1* methylation, dMMR-MLH1me; Lynch syndrome, dMMR-LS; primary *MLH1* epimutation, dMMR-PriEpi; single somatic mutation, dMMR-SS.
